# Rapid DNA methylation-based classification of pediatric brain tumours from ultrasonic aspirate specimens

**DOI:** 10.1101/2023.10.25.23297418

**Authors:** Michèle Simon, Luis P. Kuschel, Katja von Hoff, Dongsheng Yuan, Pablo Hernáiz Driever, Elisabeth G. Hain, Arend Koch, David Capper, Matthias Schulz, Ulrich-Wilhelm Thomale, Philipp Euskirchen

## Abstract

**Background:** Although cavitating ultrasonic aspirators are commonly used in neurosurgical procedures, the suitability of ultrasonic aspirator-derived tumor material for diagnostic procedures is still controversial. Here, we explore the feasibility of using ultrasonic aspirator-resected tumor tissue to classify otherwise discarded sample material by fast DNA methylation-based analysis using low pass nanopore whole genome sequencing.

**Methods:** ultrasonic aspirator-derived specimens from pediatric patients undergoing brain tumour resection were subjected to low-pass nanopore whole genome sequencing. DNA methylation-based classification using a neural network classifier and copy number variation analysis were performed. Tumor purity was estimated from copy number profiles. Results were compared to microarray (EPIC)-based routine neuropathological histomorphological and molecular evaluation.

**Results:** 18 samples with confirmed neuropathological diagnosis were evaluated. All samples were successfully sequenced and passed quality control for further analysis. DNA and sequencing characteristics from ultrasonic aspirator-derived specimens were comparable to routinely processed tumor tissue. Classification of both methods was concordant regarding methylation class in 16/18 (89%) cases. Application of a platform-specific threshold for nanopore-based classification ensured a specificity of 100%, whereas sensitivity was 78%. Copy number variation profiles were generated for all cases and matched EPIC results in 16/18 (89%) samples, even allowing the identification of diagnostically or therapeutically relevant genomic alterations.

**Conclusion:** Methylation-based classification of pediatric CNS tumors based on ultrasonic aspirator-reduced and otherwise discarded tissue is feasible using time- and cost-efficient nanopore sequencing.

## Introduction

Ultrasonic aspirator devices are frequently used in pediatric neurosurgery for efficient microsurgical resection of brain tumours while minimizing tissue damage to surrounding healthy brain (1). With ultrasonic aspirator, tumor tissue is fragmented in situ by ultrasound-induced vibration and tissue debris is aspirated using suction. To date, ultrasonic aspirator tissue specimens have not been used for routine neuropathological examinations. At the same time, molecular profiling is increasingly used and required in addition to histomorphology for diagnostic workup and comprehensive characterization of pediatric brain tumours (2). In particular molecular classification based on DNA methylation signatures has proven to be a powerful and elegant unbiased approach to identifying tumor type (3) and has been adopted in the current World Health Organization (WHO) classification of central nervous system (CNS) tumours (4). For DNA extraction, however, additional tissue is needed which may be scarce in pediatric neurosurgery. While for histological examination it appears necessary to preserve the integrity of the tissue, DNA methylation profiling (as any nucleic acid-based method) only relies on the integrity of tumor DNA. Repurposing ultrasonic aspirator tissue specimens as a source of tumor DNA for molecular diagnostics would maximize use of tumor tissue. To date, only detection of focal amplifications (5) and gene expression profiling by RNA sequencing (6) in ultrasonic aspirator tissue samples have been explored.

The growth patterns of pediatric brain tumors differ from those of adult tumors in that they are more likely to spread in the neuraxis (7). Furthermore, highly aggressive rare embryonal and sarcomatous pediatric CNS tumors for which there are limited therapeutic recommendations and for which immediate initiation of therapy is essential have only recently been molecularly redefined (8). The overall time to integrated diagnosis in pediatric oncology is therefore of considerable importance, and any delay in initiating first-line therapy may be critical. Indeed, the presence of molecular markers defining risk groups in therapy trials also leads to different therapeutic approaches.

Recently, we have demonstrated, that low-pass nanopore whole genome sequencing (WGS) is non-inferior to microarray-based DNA methylation profiling of CNS tumors (9). In addition, real-time analysis is feasible, enabling a reliable intraoperative diagnosis within a surgically relevant time frame at low cost (10). In addition, adaptive sequencing allows to enrich genomic regions of interest in the same WGS run to detect clinically relevant SNV and SV (11).

In the present study, we studied whether DNA methylation-based classification can be reliably performed using DNA from tumor tissue fragments obtained by ultrasonic aspirator devices using low-pass nanopore whole genome sequencing in order to overcome time-consuming tissue processing and maximize use of limited material in pediatric neuro-oncology.

## Methods

### Study design

We conducted a prospective, proof-of-concept single-center study using ultrasonic aspirator tissue specimens for molecular characterization of pediatric CNS tumors using nanopore WGS (Fig. 1 (A)). All patients < 18 years who underwent surgery for tumor resection using an ultrasonic aspiration device at the Department of Pediatric Neurosurgery, Charité-Universitätsmedizin Berlin, Germany, between February 6th, 2020, and October 5th, 2020, were screened. Informed written consent was obtained from patients and/or guardians. The study was approved by the local ethics committee (Charité –Universitätsmedizin Berlin, Berlin, Germany; EA2/041/18) and performed according to the guidelines for Good Scientific Practice. Ultrasonic aspirator tissue samples taken with the LEVICS device (Söring, Quickborn, Germany), which are normally discarded after surgery, are collected using a bronchoalveolar lavage trap, which is connected to the suction tubing coming from the sonotrode instruments and connected to the suction reservoir. Thereby about 5ml of fluid including fragmented tumor tissue could be collected. In parallel, regularly resected umor tissue was processed for routine neuropathological procedures including phenotypic-genotypic diagnostics. All study data were archived under an ID key accessible only to the research group (pseudonymization). Pseudonymized study data were recorded using REDCap (13), which was provided by the Berlin Institute of Health’s Clinical Research Unit in a certified computing environment.

**Figure 1:**
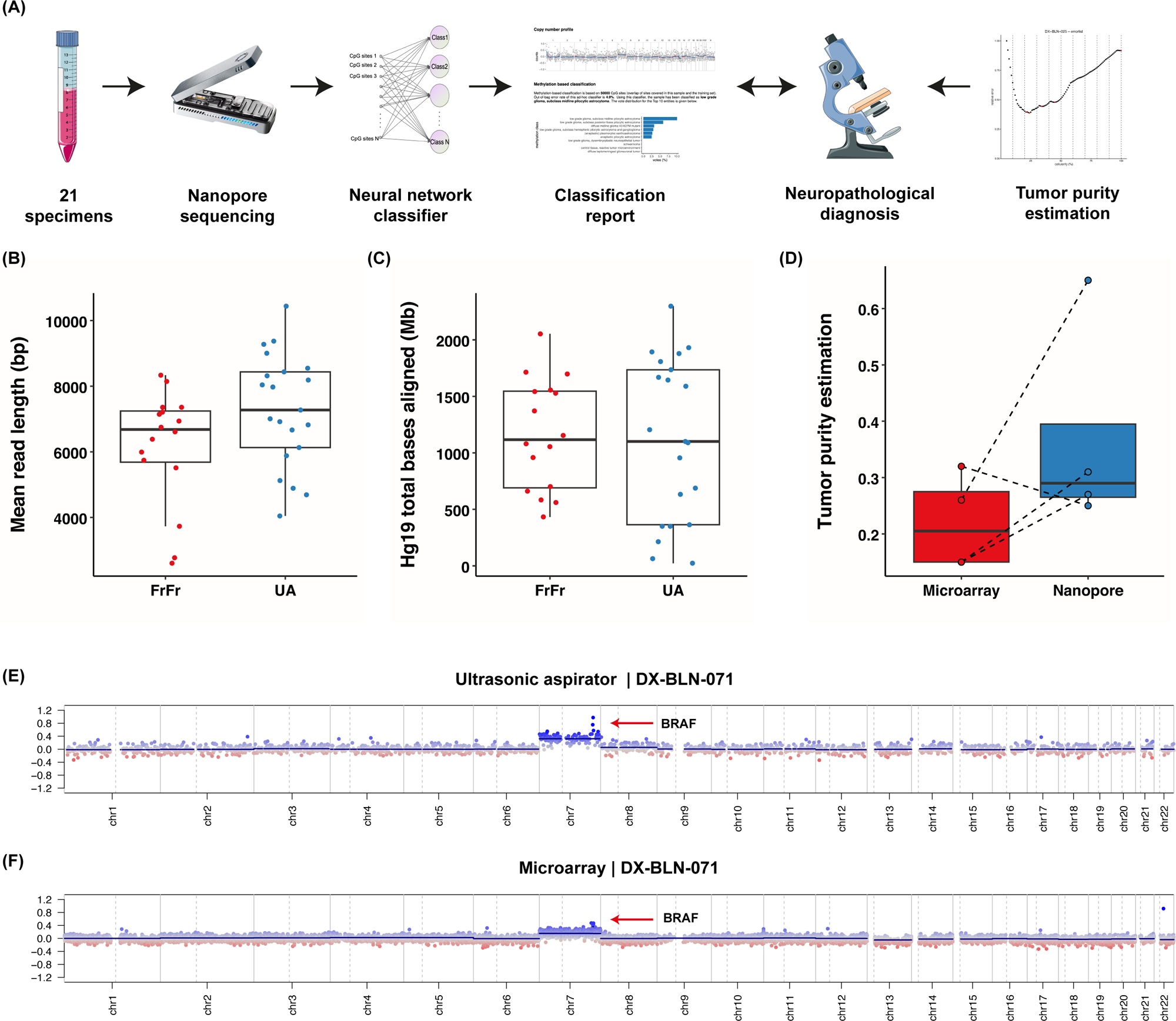
**(A)** Overview of the study design including workflow using ultrasonic aspirator tissue specimens for nanopore sequencing for DNA methylation-based classification using a neural network classifier and copy number variation analysis, comparison to microarray-based routine neuropathological profiling and assessment of tumor purity by absolute copy number estimation using ACE. Suitability of ultrasonic aspirator-derived tumor tissue (UA) for nanopore sequencing (T-Test and Mann-Whitney *U* with *P* > .05): **(B,C)** Comparison of **(B)** mean read length and **(C)** read yield obtained from standard nanopore protocol using fresh-frozen (FrFr) tumor tissue vs. ultrasonic aspirator-derived sample material indicates similar sequencing performance. **(D)** In silico tumor purity estimation between nanopore ultrasonic aspirator tissue samples and microarray FFPE tissue. **(E,F)** Representative illustration of matched copy number variation profiles obtained from **(E)** ultrasonic aspirator tissue samples and nanopore sequencing and **(F)** FFPE tumor tissue subjected to EPIC microarray (850K). Red marker indicates a low-level gain of the *BRAF* locus suggestive of a *BRAF* gene fusion.

### Ultrasonic aspirator tissue sample processing

Fresh ultrasonic aspiratior fluid aliquots were centrifuged at 1.000 rpm for 5 min,supernatant was discarded and pellets stored at -40°C. DNA was extracted from ∼25 mg thawed aspirate and purified using the DNeasy Blood & Tissue Kit (Qiagen, NL). Based on the 260/280 ratio (NanoDrop, Thermo Fisher, USA), DNA quality was determined, followed by DNA quantification with a Qubit 4.0 fluorometer using a dsDNA BR Assay (Thermo Fisher, USA).

### Nanopore low-pass whole genome sequencing

All samples were subjected to low-pass whole genome sequencing as described previously (9). Preprocessing of raw data for sequencing was performed using the publicly available nanoDx pipeline (https://gitlab.com/pesk/nanoDx, v0.5.1). Briefly, basecalling of nanopore FAST5 raw data was performed using guppy v5.0.16 (Oxford Nanopore Technologies, UK) and aligned to the hg19 human reference genome using minimap2 v2.15 (14). In order to assess the feasibility of ultrasonic aspirator-derived nanopore sequencing, the aligned sequencing data was normalized to a six hour sequencing window and were compared to a previously published dataset of 16 brain tumor samples obtained during routine nanopore sequencing from fresh-frozen tumor tissue (9). DNA methylation was assessed using nanopolish v0.11.1 (15). After binarization of beta values with threshold = 0.6 (9), features with zero variance were filtered out, leading to 366,263 CpG sites retained. These were used to train the neural network model with randomly masked features. PyTorch, an open source deep-learning framework, was used to develop the model (16). To obtain class probability estimates that can be used to guide diagnostic decision-making, a normalization function and a Softmax layer was used to convert the raw values into a probability that measures the confidence in the brain tumor class assignment (the calibrated score). Returned majority votes were combined to methylation class families (MCF), if feasible (9). Complementarily, data was displayed using t-distributed Stochastic Neighborhood Embedding (t-SNE) (17).

### CNS tumor classification

All cases were classified according to the 2016 WHO CNS classification during routine neuropathological examination at the Department of Neuropathology, Charité Universitätsmedizin Berlin. The recent 2021 edition (4) was not yet available during the study period of this patient cohort. Nanopore classification results were compared to the reference diagnosis as well as microarray-based classification results of the same tumor considering the established cut-off values for the probability score.

### Copy number analysis

Copy number variation (CNV) analysis from nanopore WGS data was performed using the QDNAseq package v1.8.0 and R/Bioconductor v3.3 as described before (18, 19). To account for region- and technology-specific artifacts, public nanopore WGS data for the NA12878 human reference genome were processed and subtracted from the normalized bin counts of the tumor samples for case reports. To estimate tumor purity in aneuploid tumors, absolute copy number estimation of nanopore- and microarray-based data was performed using the ACE software package (v1.6.0) (20). All estimates were verified manually.

### Methylation array processing

Infinium MethylationEPIC BeadChip microarrays (Illumina) were used to obtain genome-wide DNA methylation profiles for tumor samples during routine neuropathological diagnostic examination. Data were generated following the manufacturer’s protocol at the Department of Neuropathology, Charité - Universitätsmedizin Berlin, using >250 ng of DNA from FFPE tissues as input material. For classification, IDAT files were uploaded to the public Heidelberg brain tumour classifier available at https://www.molecularneuropathology.org (v.11b4).

### Statistical analysis

Reproducible and easy-to-deploy pipelines were implemented using snakemake (v.7.15.2) (21). Data analysis was mainly performed using R (v.4.0.2). Figures were designed using ggplot2 (v.3.3.2). Statistical analyses were performed using IBM SPSS^®^ 29 (Armonk, N.Y., USA)

### Data and code availability

The nanoDx analysis pipeline for end-to-end analysis of nanopore WGS data is available at https://gitlab.com/pesk/nanoDx (v.0.5.1). Source code to reproduce all analyses and sequencing data based figures in this work is provided at https://gitlab.com/pesk/nanoCUSA. Raw sequencing data have been deposited at the European Genome-phenome archive (accession no. tbd), while methylation microarray data and nanopore methylation calls are available from Gene Omnibus Express (accession no. tbd).

## Results

### Patient characteristics

A total of 21 children undergoing surgery participated in the study. 3/21 (14.9%) of patients were excluded from analysis due to non-diagnostic scores in microarray-based classification (n=2) or final diagnosis of non-neoplastic disease (n=1). Eventually, our cohort comprised 18 tumor aspirates from 18 pediatric patients (Table 1). 33% of patients (n=6) were male. Median age at surgery was 7.5 years (range 1 to 17 years). Twelve patients suffered from a newly diagnosed cerebral lesion, whereas six samples were obtained from a second or further intervention. Five patients had received previous treatment with vincristine/carboplatin according to the European guidelines for low-grade glioma (LGG) (n=4) or cyclophosphamide/vincristine/methotrexate/carboplatin/etoposide according to the current treatment recommendation of the German Society of Pediatric Oncology and Hematology (GPOH) for newly diagnosed medulloblastoma, ependymoma, and pineoblastoma (n=1). One patient with LGG was previously treated with vinblastine monotherapy and targeted therapy using a MEK inhibitor. The most frequent diagnosis was pilocytic astrocytoma (50%, n=9).

**Table 1:**
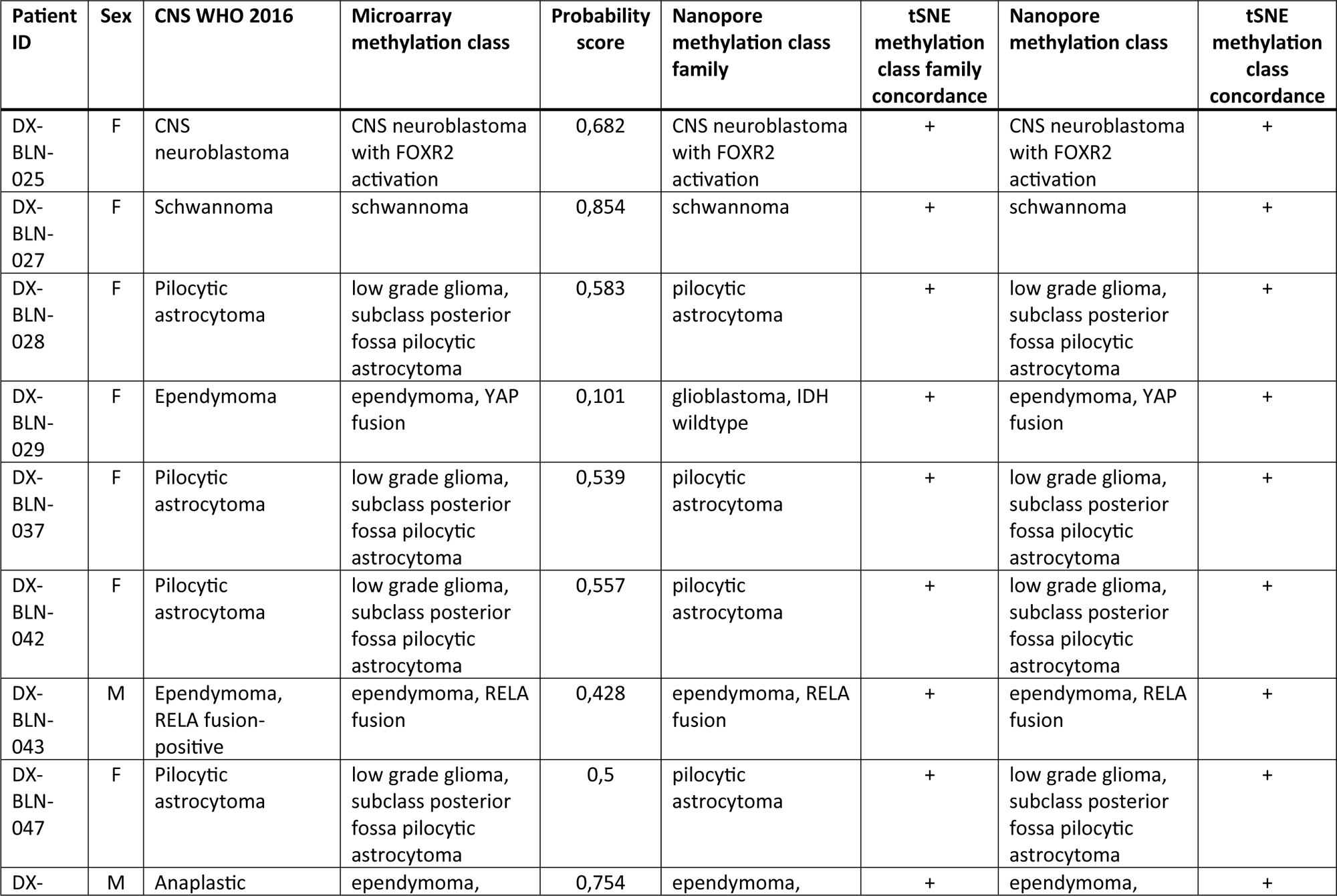
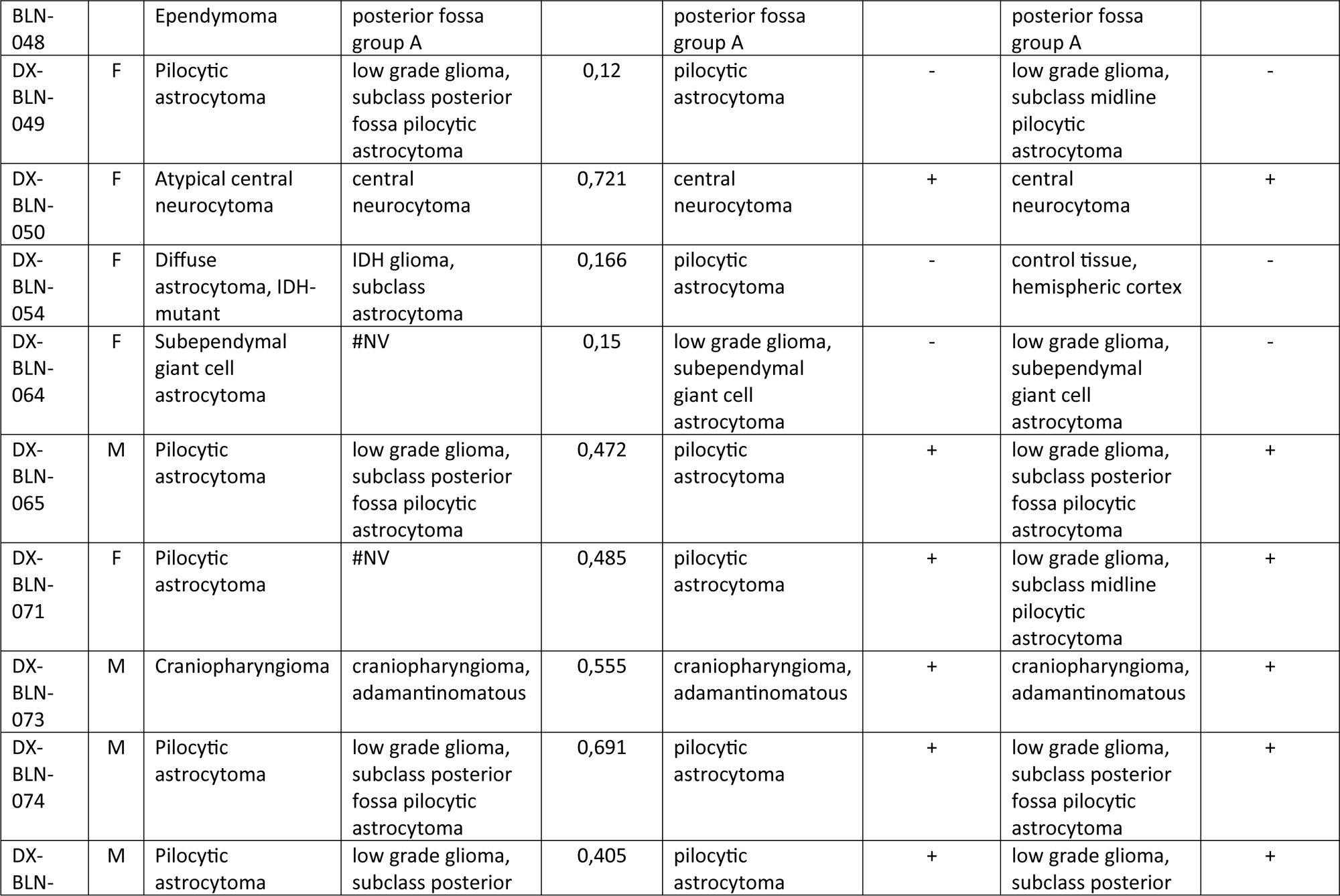
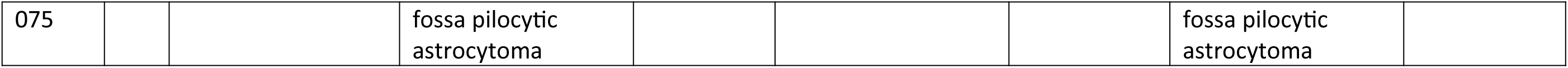
Overview of the patient cohort.

### Sequencing characteristics of tumor DNA from ultrasonic aspirator tissue samples

Low-pass whole genome sequencing performed for at least 6 hours resulted in a mean genome coverage of 0.44X (range 0.01X to 1.5X, Suppl. Table 1). The mean read length ranged between 4,047 and 10,440 base pairs with a mean of 7,377 base pairs and was comparable to reads obtained in an external cohort of sequencing runs of tumor DNA extracted from fresh-frozen tissue (Fig. 1B). The mean number of CpG sites overlapping the reference atlas was 100,852 CpGs (range: 2,275 to 295,872 CpG sites), exceeding the minimum number of 1,000 overlapping CpG sites for meaningful analysis in 18/18 (100%) samples. In two cases, the minimum number of CpG sites was not achieved initially and required an additional sequencing run. On average, after six hours of sequencing 1,054 Mb of aligned base pairs were obtained (range: 23.76 Mb - 2299.12 Mb), which again was comparable to sequencing runs from fresh-frozen tissue (Fig. 1C). Tumor cell content was reliably estimated from copy number alterations in 4/20 (20%) tumors with a mean tumor purity of 0.37 (range 0.25 – 0.65) (Fig. 1D). Tumor purity was higher in ultrasonic aspirator tissue samples compared to FFPE tissue for routine workup in 3 out of 4 (75%) cases.

### DNA methylation-based classification

Tumors were classified based on DNA methylation profiles using a neural network model which had been trained using the Heidelberg brain tumour reference cohort with CNS tumor methylation datasets of 2,801 samples and predictions were made with respect to the 91 methylation classes (MC) or methylation class family (MCF), respectively, as defined in the 11b4 version (3). A single nanopore-specific cut-off value was determined by recalibrating the raw values to identify valid predictions. Receiver operating characteristic curve analysis of the maximum calibrated scores was used to determine an optimal cut-off value > 0.2.

Classification results were identical to microarray in 16/18 (89%) of cases and compatible with the integrative histopathological reference diagnosis in 16/18 (89%) cases (Figure 2). Application of the optimal calibrated score threshold of >0.2 resulted in 14/18 cases passing the cut-off all of which were correctly classified, corresponding to a specificity of 100% and a sensitivity of 78% on both MC and MCF level (Figure 2).

**Figure 2:**
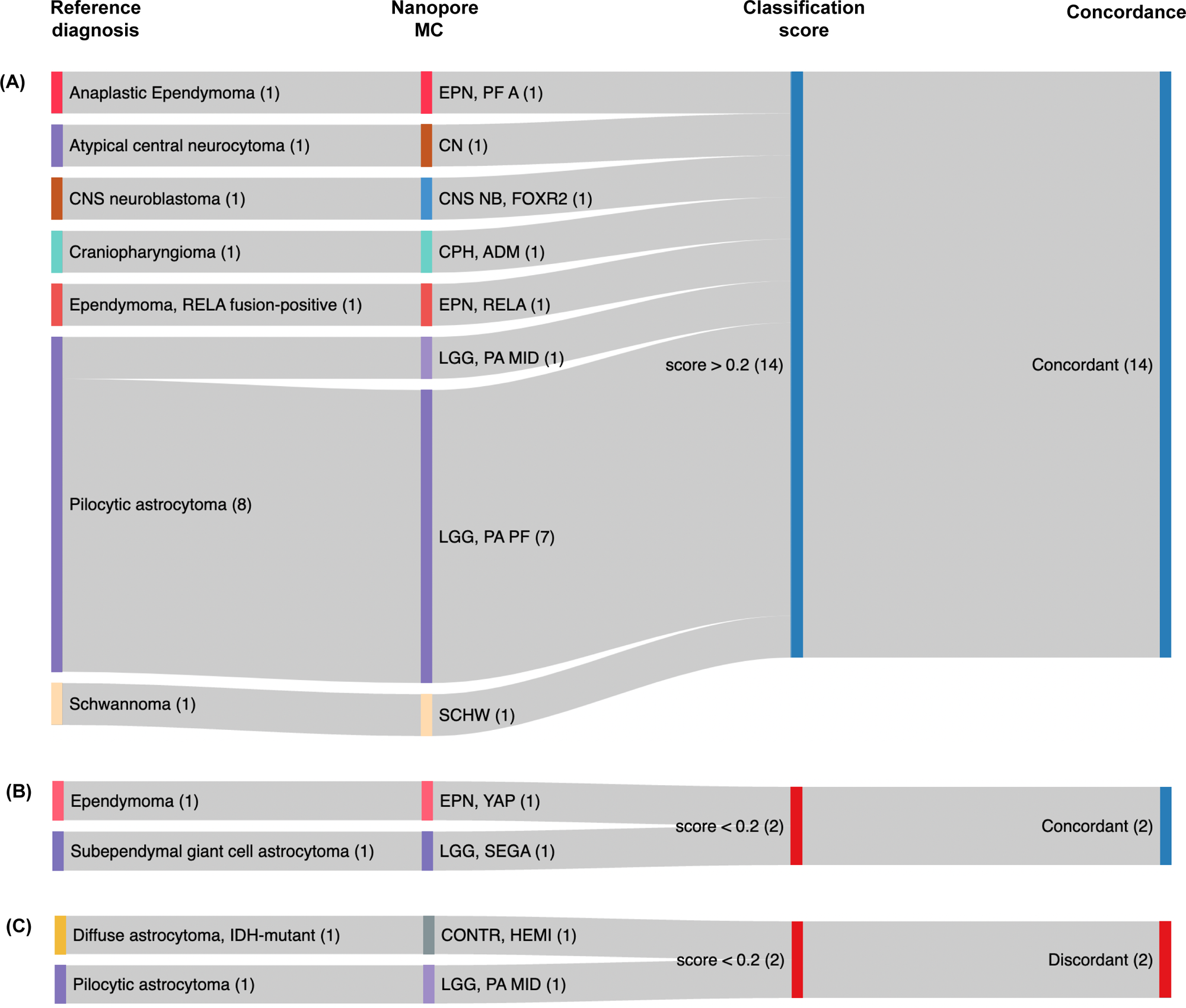
Nanopore classification methylation class call of ultrasonic aspirator-derived tumor material and the corresponding 2016 WHO CNS reference diagnosis results showing a specificity of 100% and sensitivity of 77.7 % for cases above the calibrated threshold of > 0.2.

At the MC level, in 2/18 (11%) cases, the score was below the optimal threshold but classification was still correct (one YAP-fusion positive ependymoma, one subependymal giant-cell astrocytoma (SEGA)). 2/18 (11%) cases had discordant results with scores that were below the calibrated nanopore-specific threshold: In the first case, pilocytic astrocytoma (PA) subtype was incorrect (classifying the case as midline PA instead of posterior fossa PA) while correctly classifying the sample as PA on MCF level. The other sample was an IDH-mutant astrocytoma classified as control tissue. Here, additional Sanger sequencing revealed no IDH mutation in the ultrasonic aspirator tissue sample, potentially indicating a sampling issue.

For comparison, the microarray-based analysis from matched FFPE tissue yielded a diagnostic score in 16/18 (89%) cases. Of note, the two cases with nondiagnostic score included one case of SEGA that also received a low score in nanopore/ultrasonic aspirator tissue profiling.

### Copy number profiling

Copy number profiles obtained from WGS closely resembled matched array-based profiles in 16/18 (88.9 %) cases and enabled the detection of large chromosomal alterations, whereas two nanopore-based CNVs (both pilocytic astrocytomas) were insufficient for interpretation (Suppl. Figure 1). In contrast, low-level focal gains such as tandem duplications resulting in *BRAF* fusion genes in the PA samples were visually identified in 1/9 (11.1 %) cases from nanopore CNV profiles compared with detection in 8/9 (88.8 %) in matched CNV profiles from methylation microarrays (Suppl. Figure 1).

## Discussion

Molecular testing is an essential component of state-of-the-art integrated neuropathological diagnostics for most pediatric brain tumor types. Due to the increasing number of pathological examinations required (such as DNA and RNA gene panel sequencing or methylation microarray), tissue is valuable. This proof-of-principle study reports, to our knowledge, the first application of ultrasonic aspirator-derived tumor tissue for molecular classification of pediatric CNS tumors using low-pass nanopore whole genome sequencing. We show that ultrasonic aspirator-derived tumor fragments are a representative source of tumor DNA with tumor cell content sufficient to DNA methylation-based classification and yielding identical classification results.

### Tissue characteristics

Although the use of ultrasonically minced tumor tissue for histopathological analysis of brain tumor tissue has been demonstrated in some studies, the suitability is still matter of debate (23–26). In particular, the grading of glial tumors has been reported difficult as histomorphology was only partly recapitulated.

Using read length distribution of mapped nanopore reads as a proxy of DNA fragment length, we find no significant difference in DNA extracted from fresh ultrasonic aspirator tissue aspirates compared to routinely prepared fresh-frozen tissue. Our analysis thus confirms that high molecular weight genomic DNA suitable for nanopore sequencing can be extracted when ultrasonic aspirator-derived tissue is used. Additionally, similar aligned base yields indicate comparable sequencing performance. Tumor purity estimations for microarray and nanopore indicate a tendency towards higher tumor purity in ultrasonic aspirator tissue samples. However, the estimation depends on the existence of numerical chromosomal alterations. As expected, the majority of cases within a pediatric cohort are pilocytic astrocytomas which lack relevant aneuploidy. Therefore, tumor purity could only be determined in 4 cases.

One of the major advantages of using ultrasonic aspirator tissue samples is that multiple areas of the excised tumor are sampled (23). This is particularly important because analyses based on single biopsies may have potential consequences for treatment decisions in spatially and temporally heterogeneous pediatric tumors (27, 28). In contrast, DNA extraction for methylome profiling is usually done after microdissection of a representative area of the tumor sample with an anticipated tumor cell content of ≥70% and therefore reflects only a subset of the entire tumor (3).

### Comparison to microarray-based classification

It was recently demonstrated, that the application of nanopore technology can be used with comparable reliability for processing of fresh frozen tissue compared to microarray-based analysis of FFPE material (9). Our analysis confirms its suitability when using ultrasonic aspirator-derived tissue. Similar to the observed sensitivity of 88% in a well-defined validation cohort for microarray-based classification (3), our approach reaches an overall accuracy of 89% and a sensitity of 78% for the > 0.2 cut-off while retaining 100% specificity. In contrast, in a real-world cohort enriched for challenging cases a sensitivity as low as 56% was reported for EPIC-based microarray analysis (29).

Especially low-tumor cell content, like in the infiltration zone of diffuse glioma, can be challenging for the performance of DNA methylation-based classification (30) and was likely the cause for the discordantly classified cases (before application of diagnostic cut-offs) in this cohort. Therefore, low tumor cell content poses a challenge to methylation-based classification in general, independent of the processed tissue type or technology platform used for methylome profiling.

Copy number profiles can be derived both from nanopore low-pass WGS and methylation microarray datasets. While the resolution of microarray-based CNV plots is fixed due to the probe set of the chip, resolution of low-pass WGS-based CNV plots correlates with read yield. In our cohort, the quality of nanopore CNV plots was frequently inferior to the matched microarray-based ones. However, next generation flow cells and chemistries for nanopore sequencing devices offer better yields and are likely to resolve these issues.

#### Conclusion

DNA methylation-based classification of pediatric CNS tumors from ultrasonic aspirator-fragmented tissue is feasible using nanopore sequencing. A neural network classifier with nanopore-specific diagnostic score thresholds assures high specificity while achieving acceptable sensitivity. Generation of CN profiles is possible and allows for detection of chromosomal changes, but was currently inferior in detection of focal changes (e.g. *BRAF* tandem duplication) compared to microarray approaches. This approach allows maximum exploitation of available tissue for diagnostics. Since advanced molecular techniques have limited benefit for patients in ressource-challenged centers, our time- and cost-efficient approach may be of particular interest.

## Acknowledgments

We thank Aydah Sabah for expert technical assistance. Computation has been performed on the HPC for Research cluster of the Berlin Institute of Health. P.E. has been a participant in the BIH-Charité Clinical Scientist Program funded by the Charité – Universitätsmedizin Berlin and BIH.

Figures were created in part using Servier Medical Art provided by Servier and licensed under a Creative Commons Attribution 3.0 unported license. In addition, images illustrating nanopore sequencing were reproduced with permission from Oxford Nanopore Technologies Plc, United Kingdom.

---

**Suppl. Figure 1:**
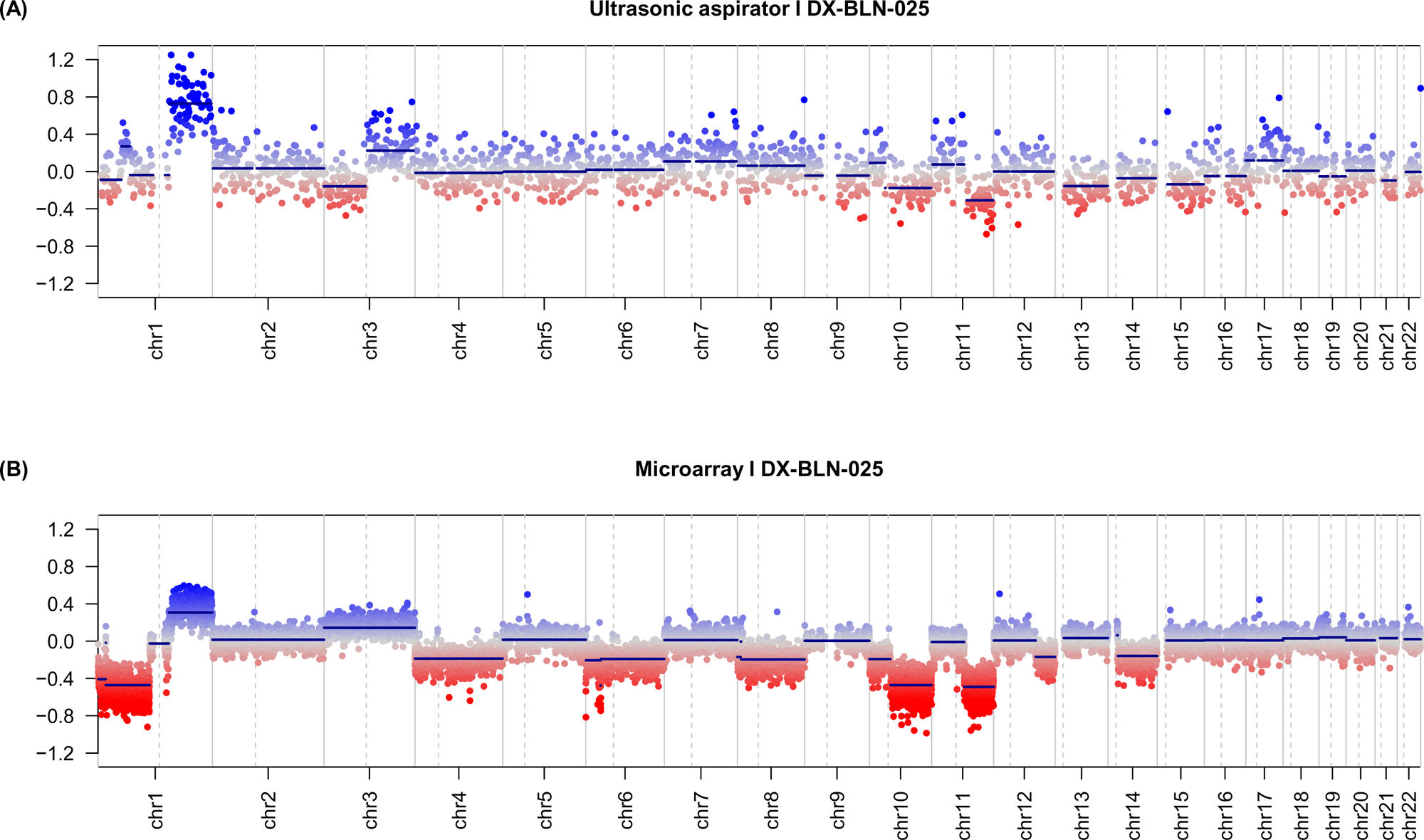
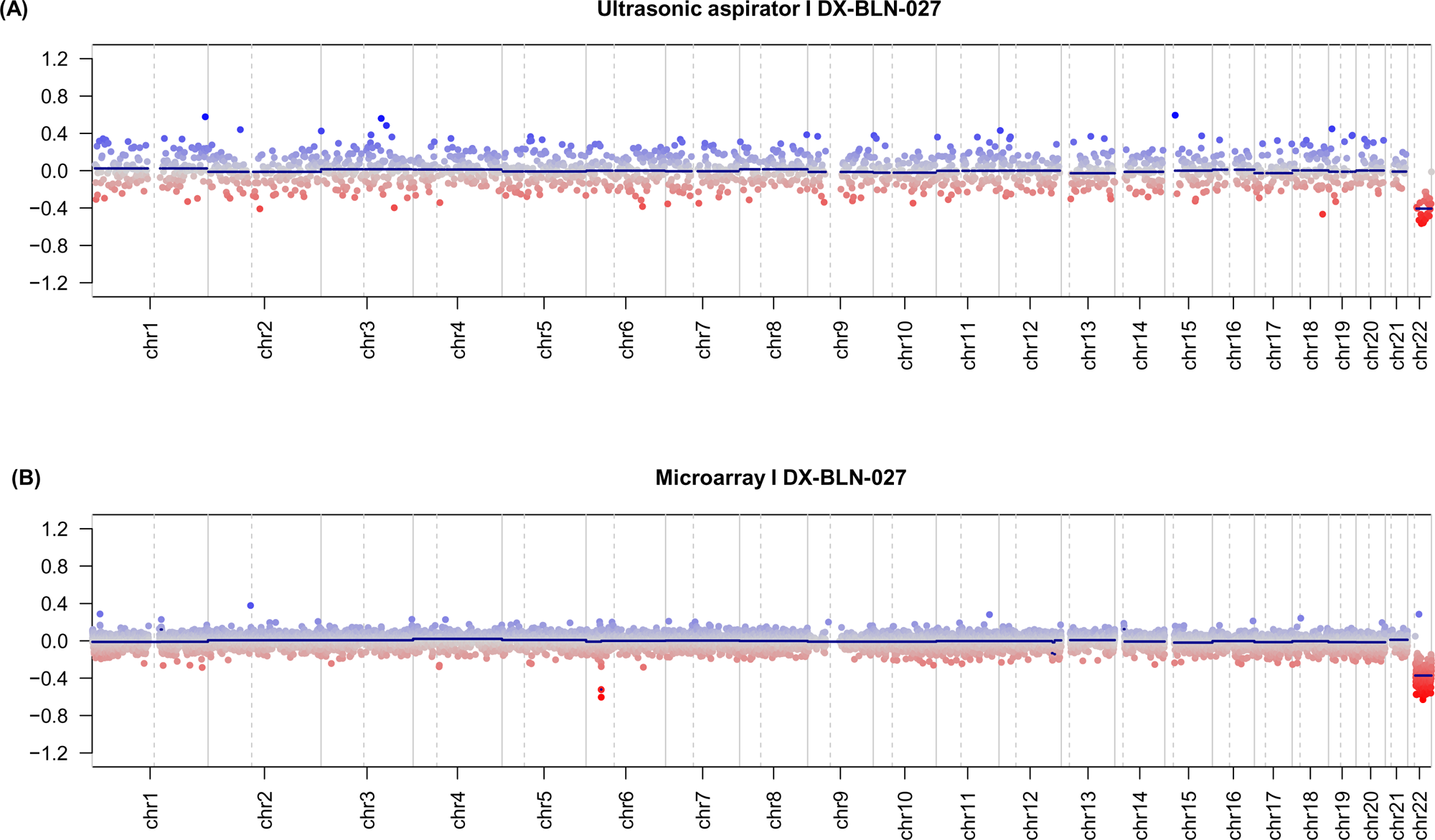
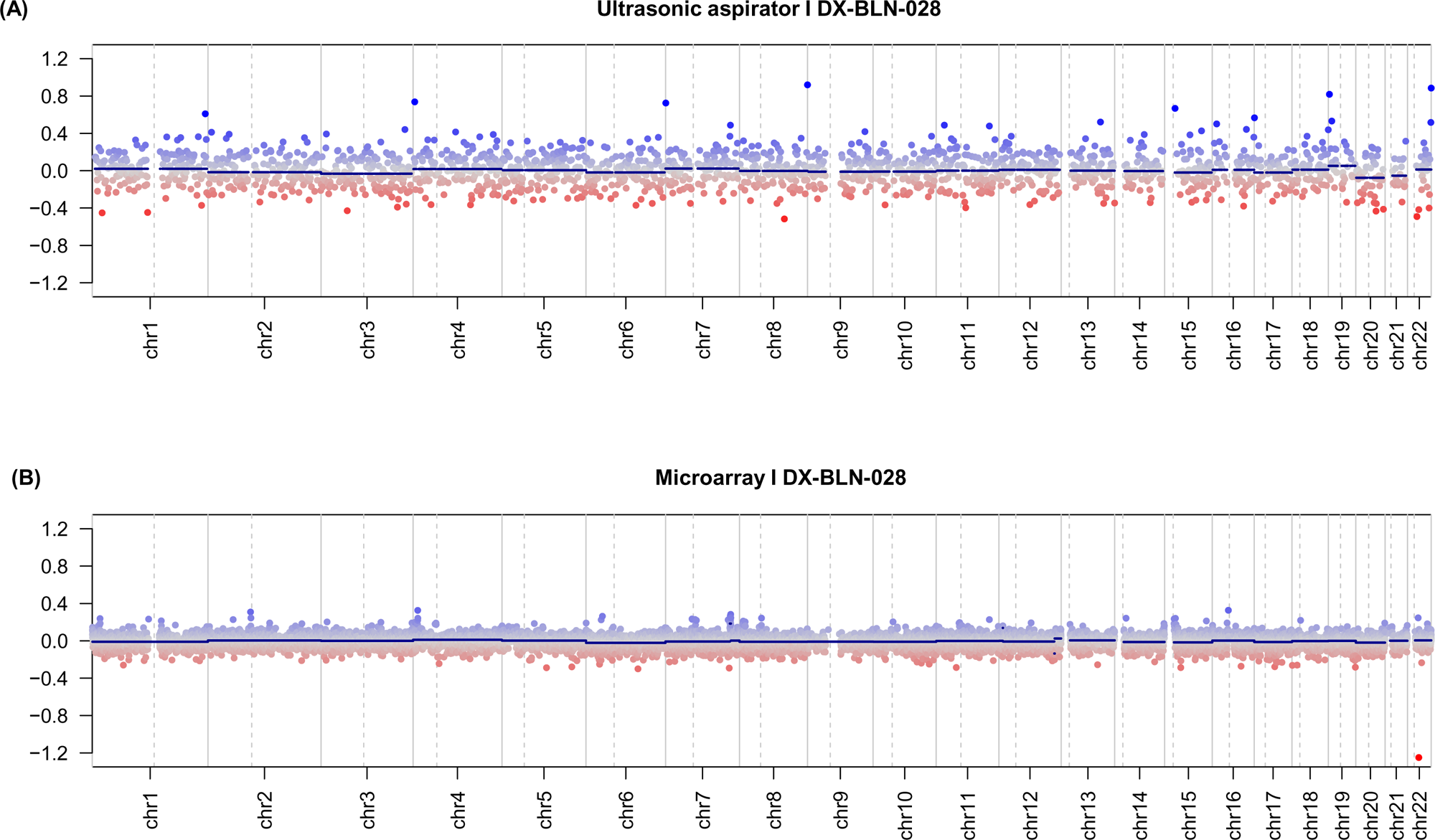
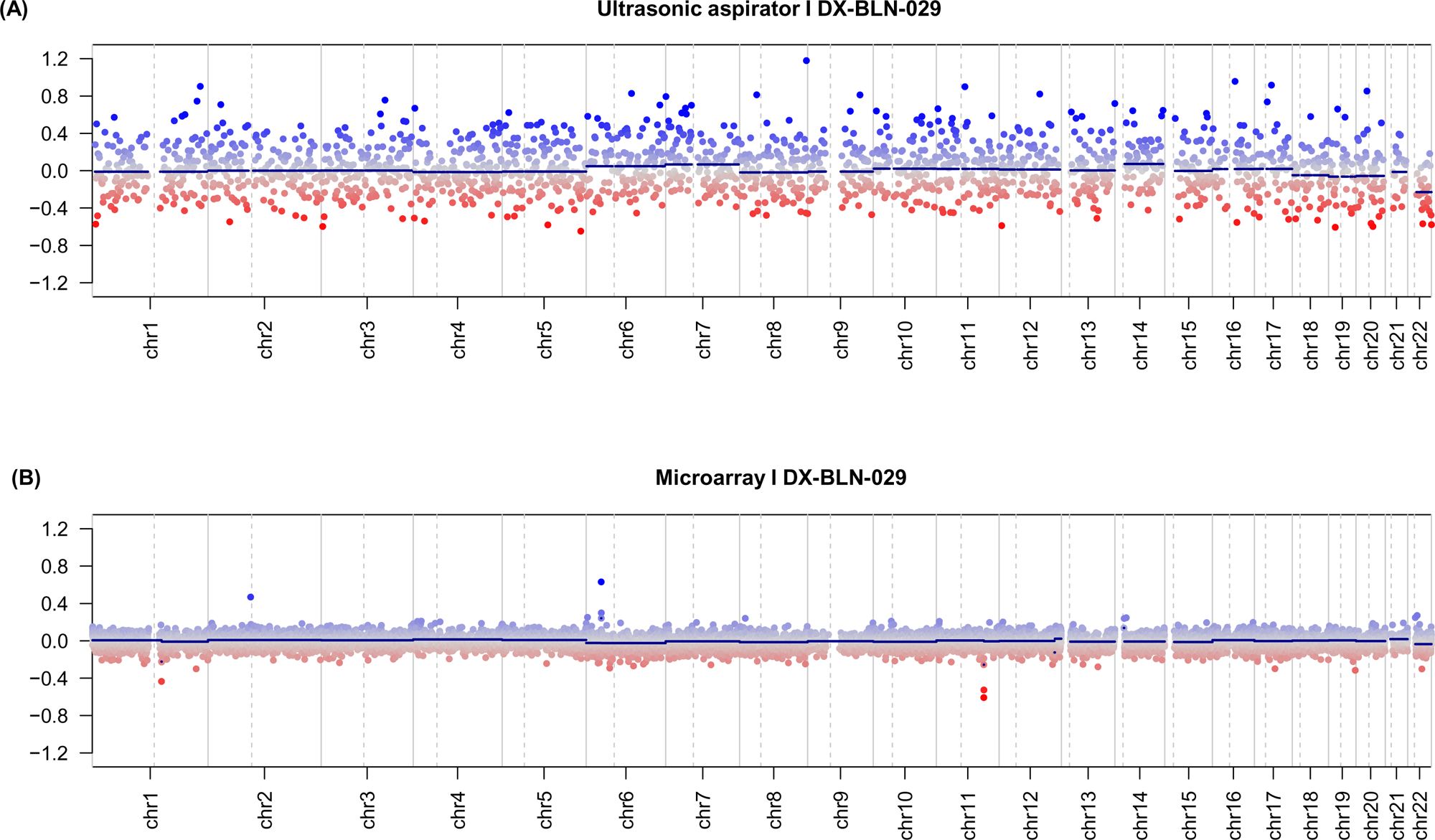
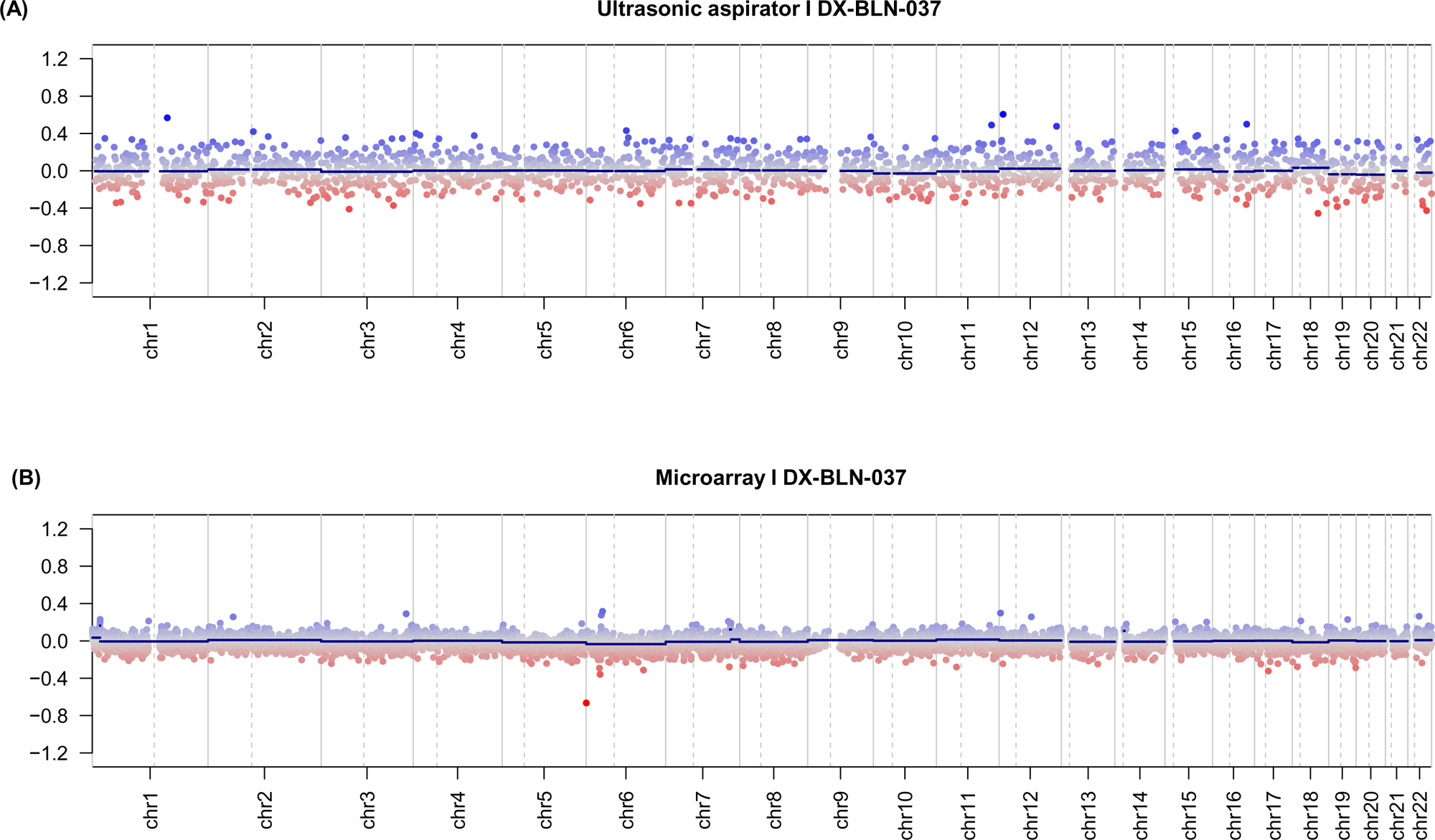
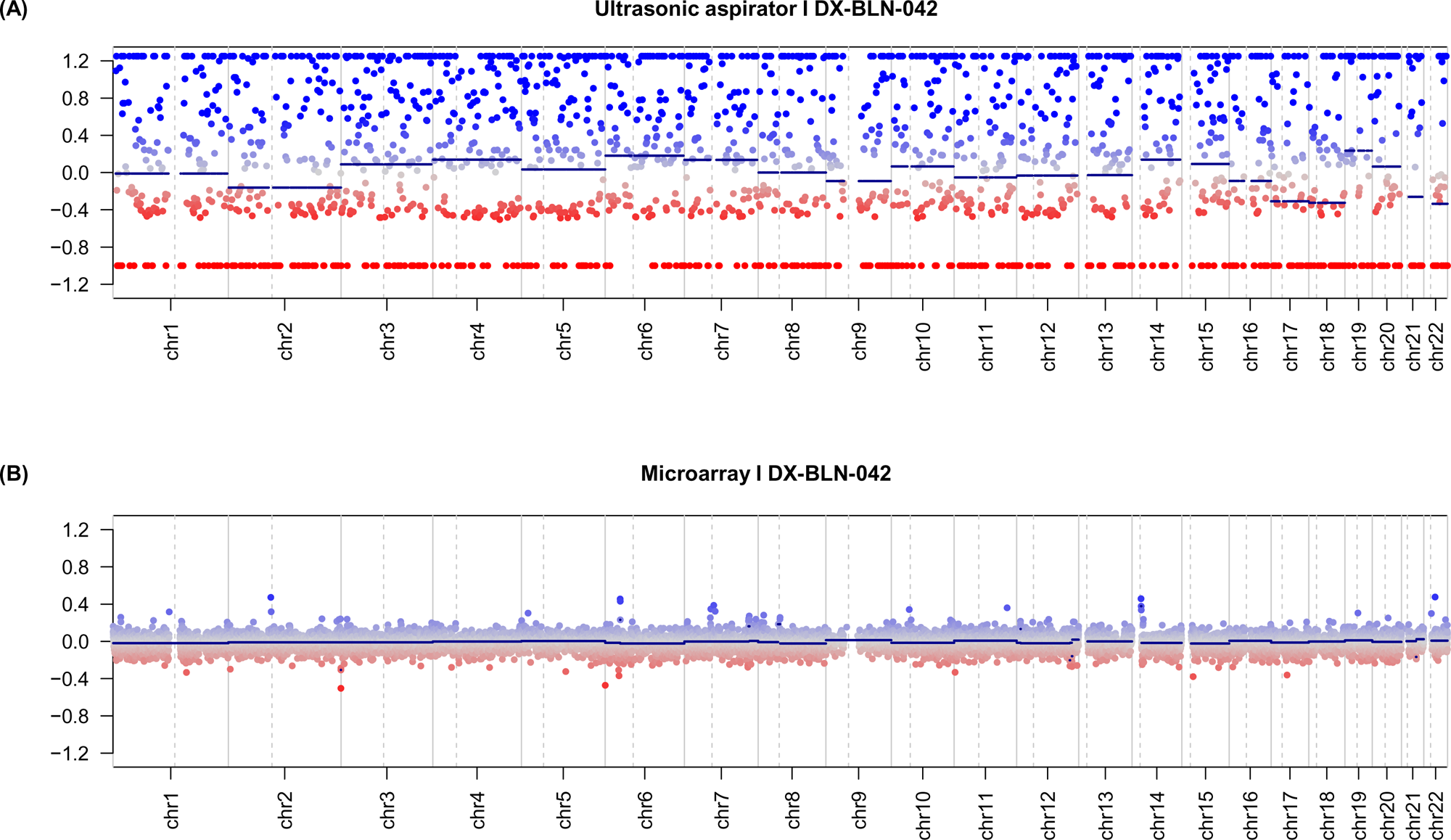
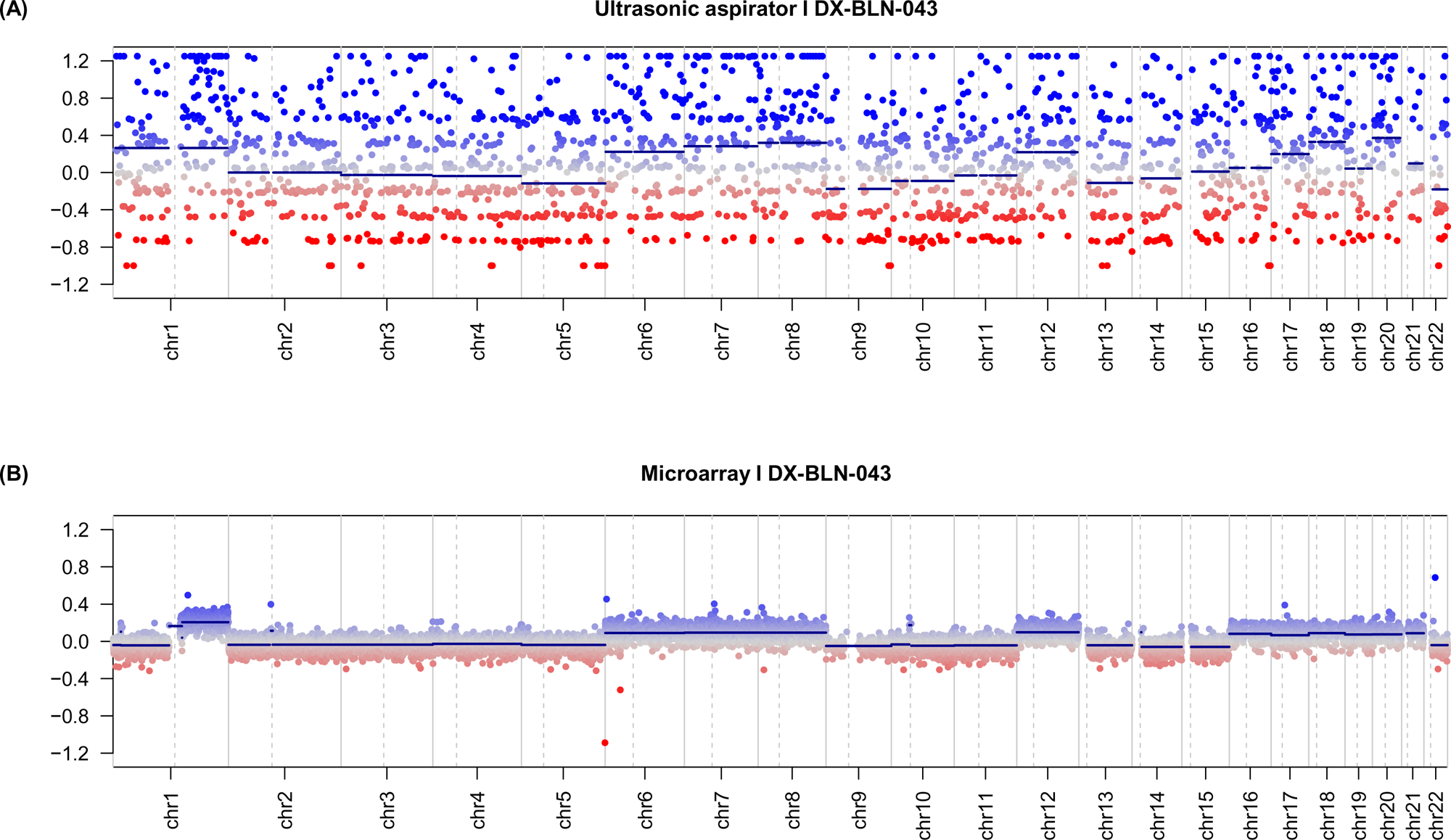
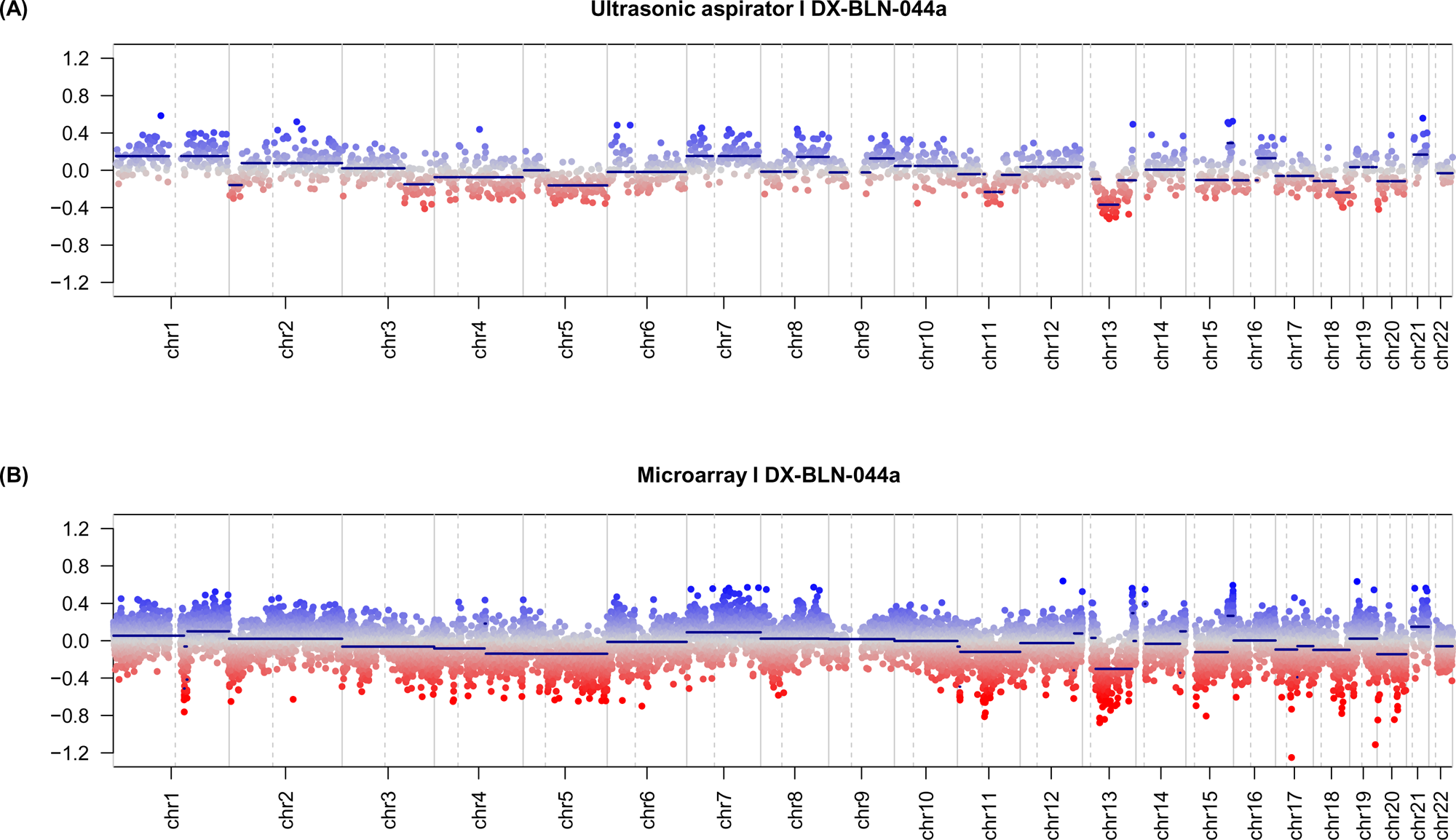
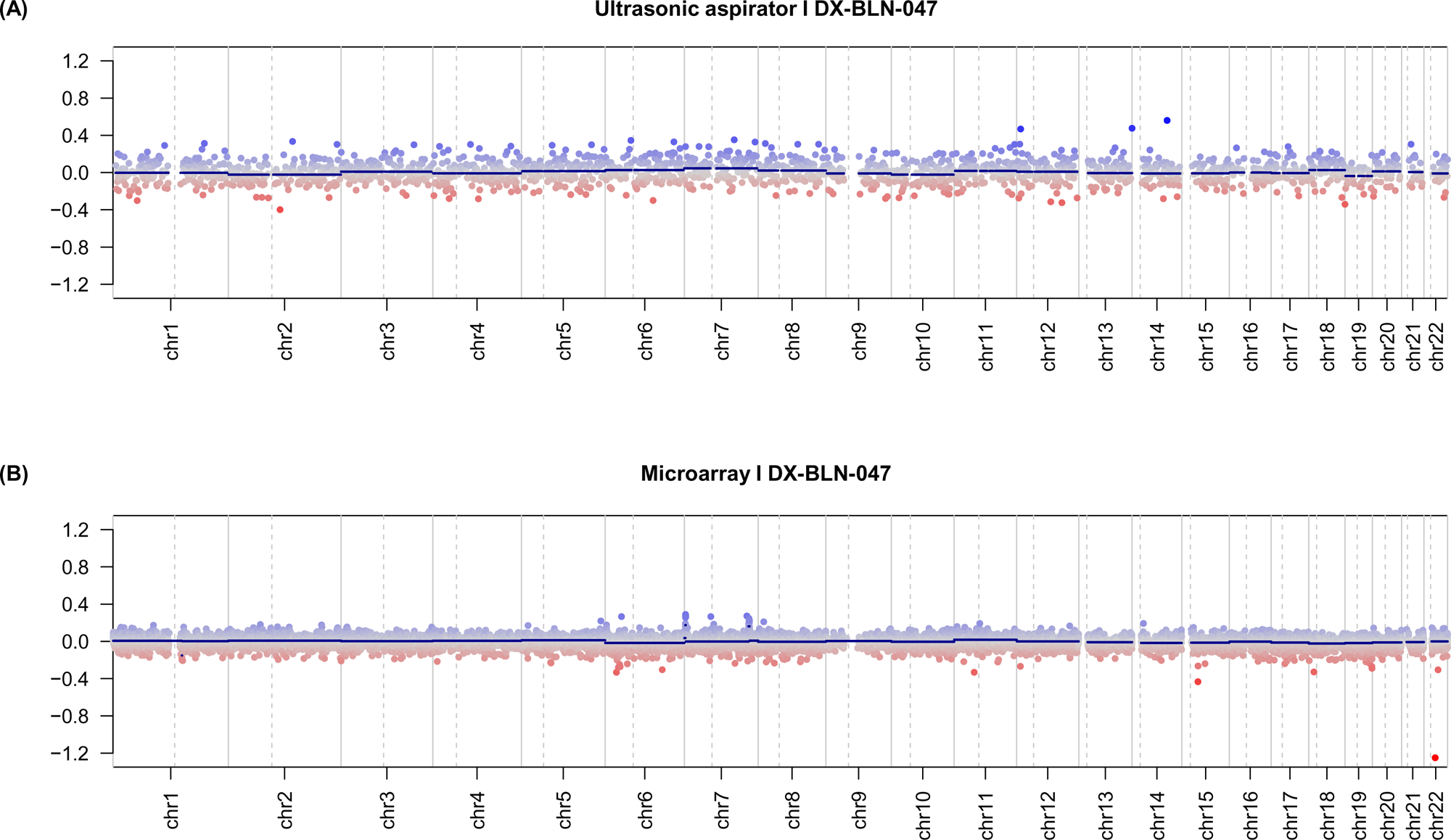
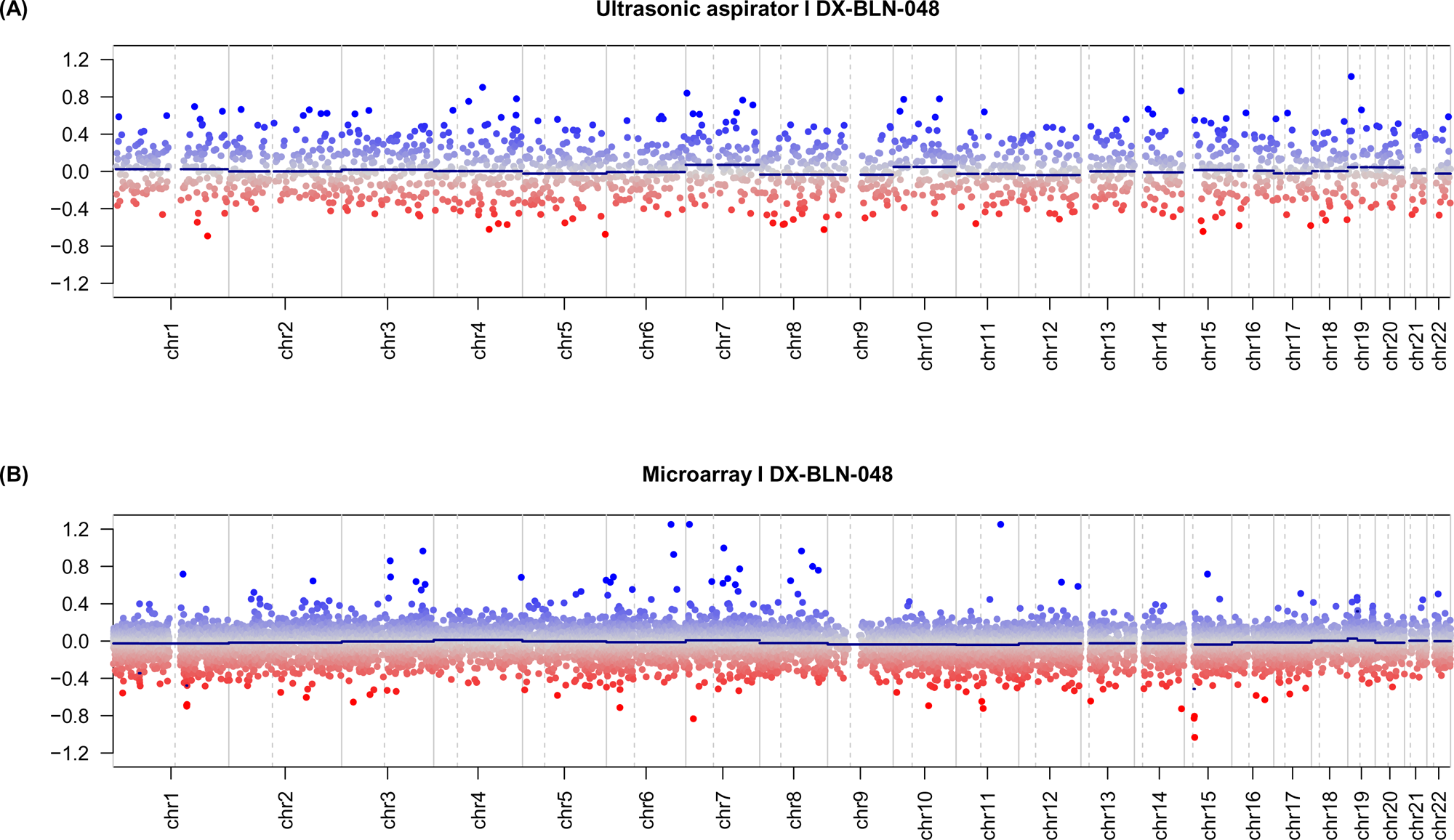
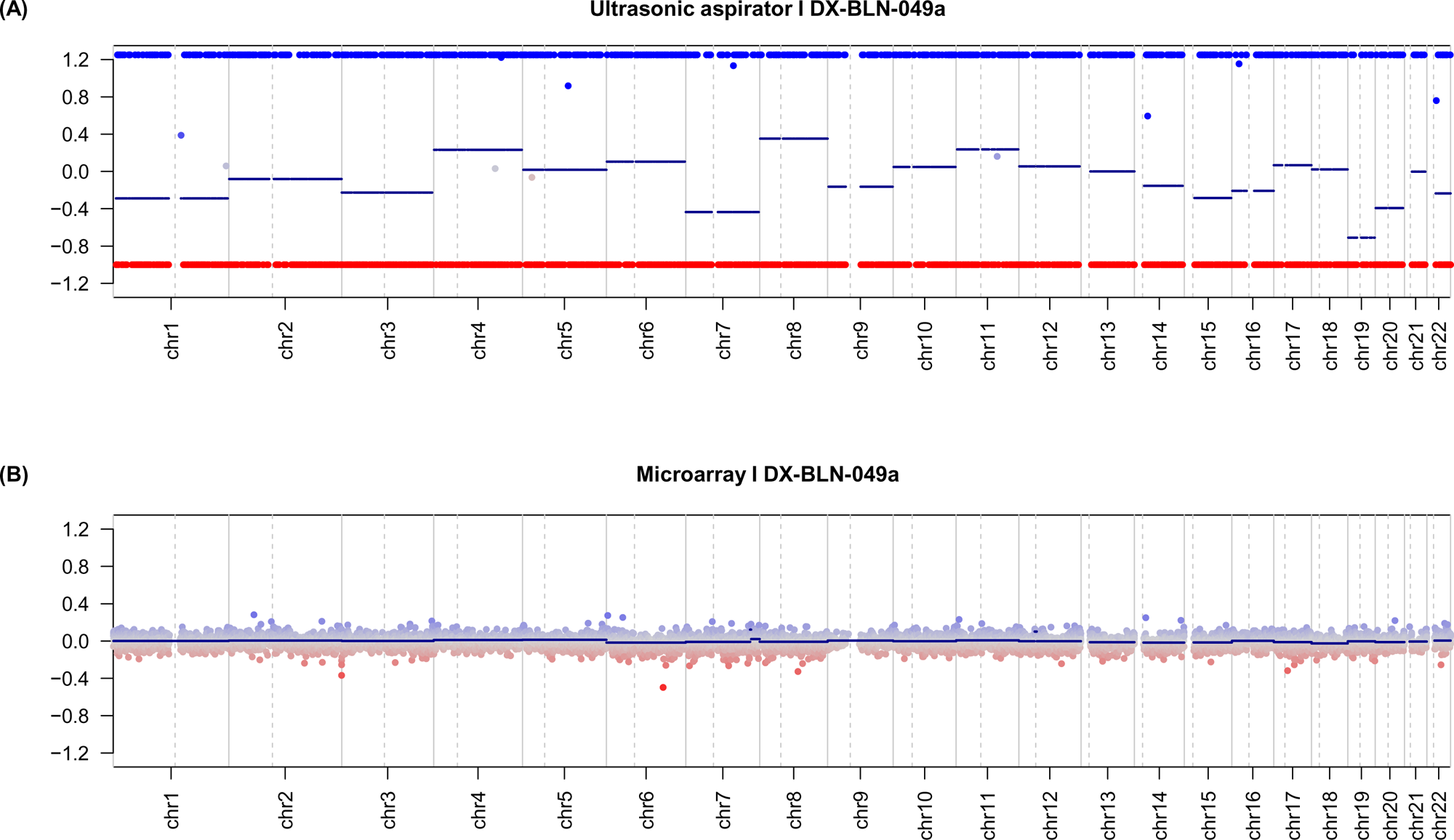
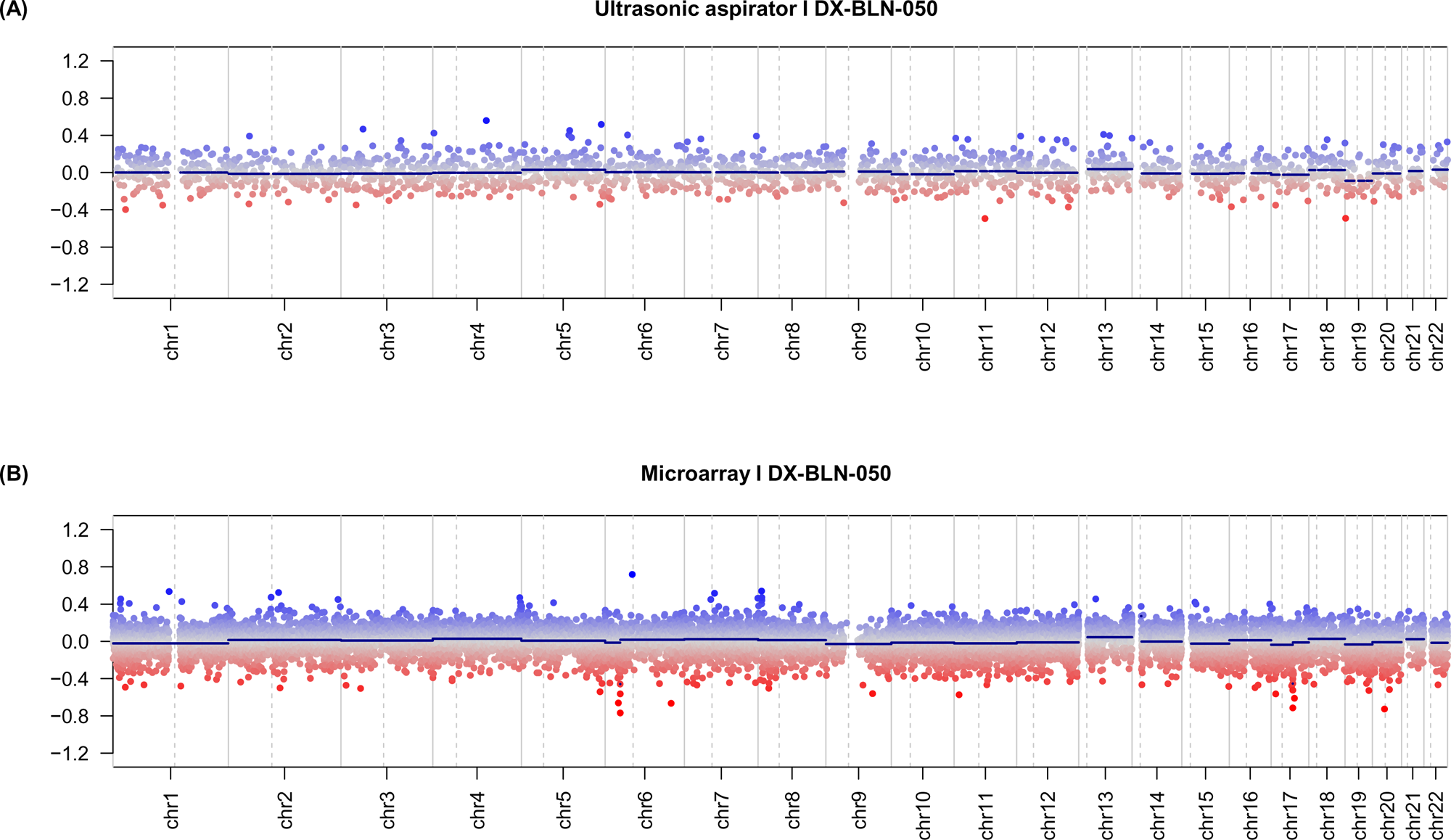
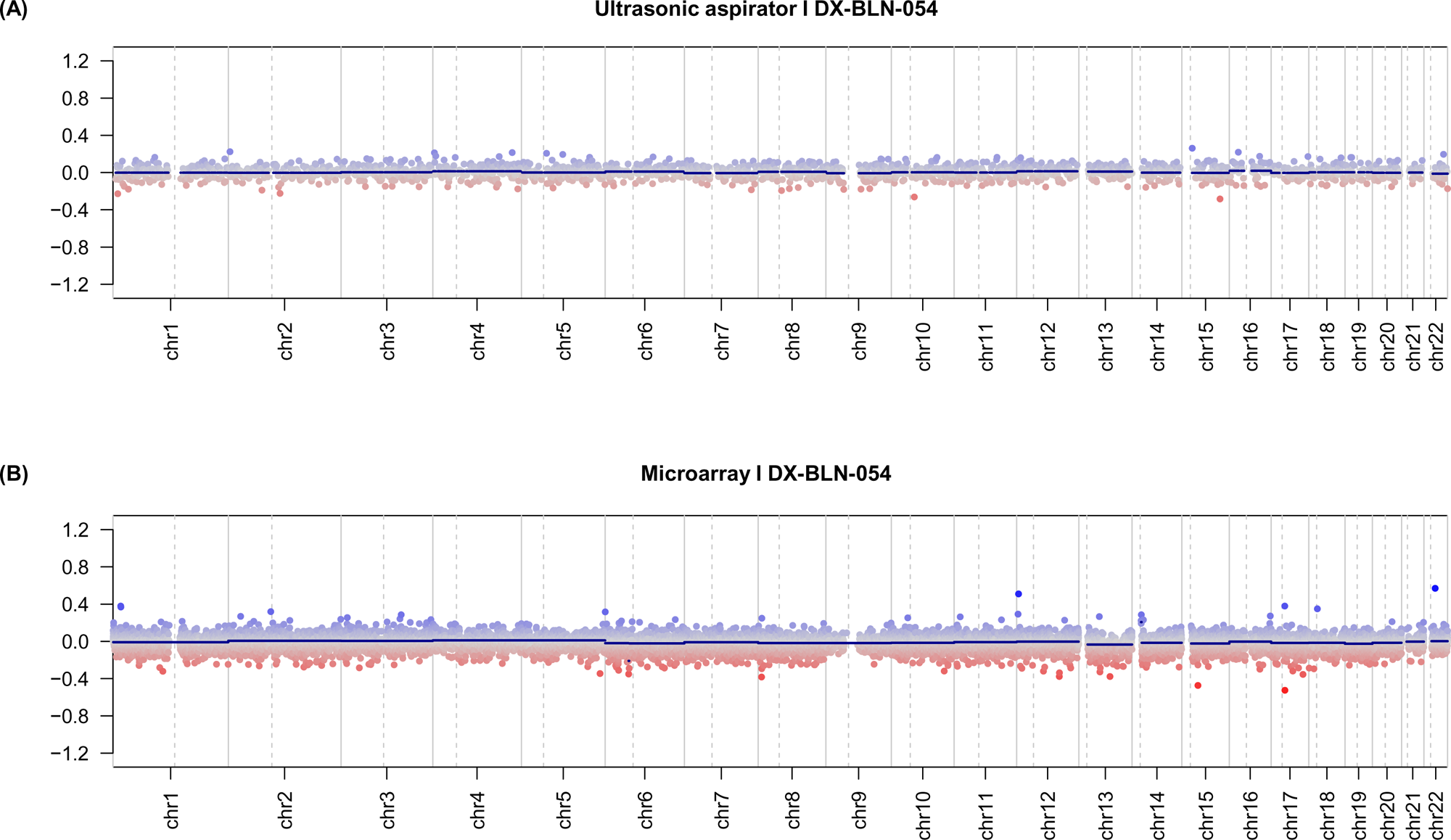
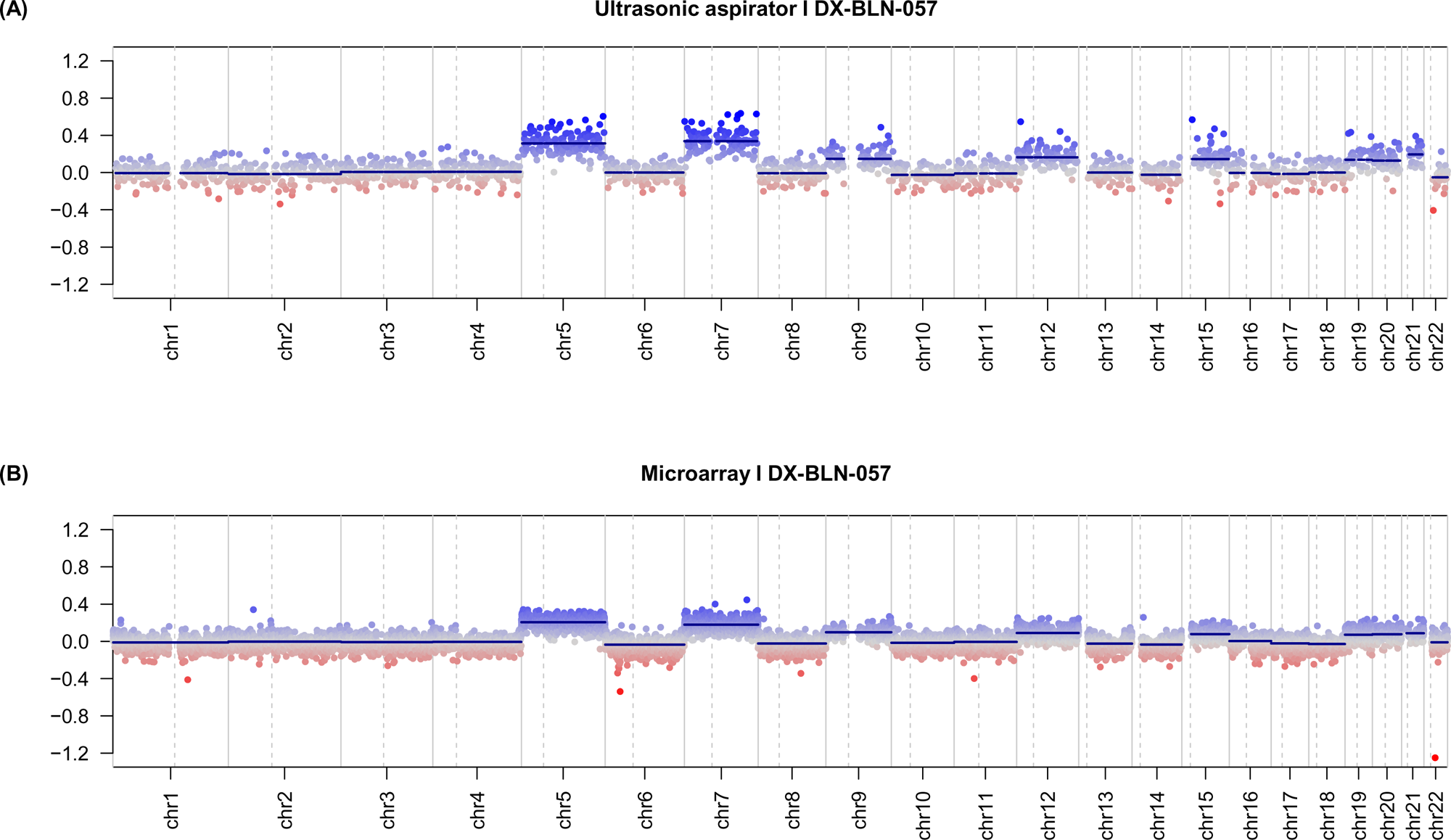
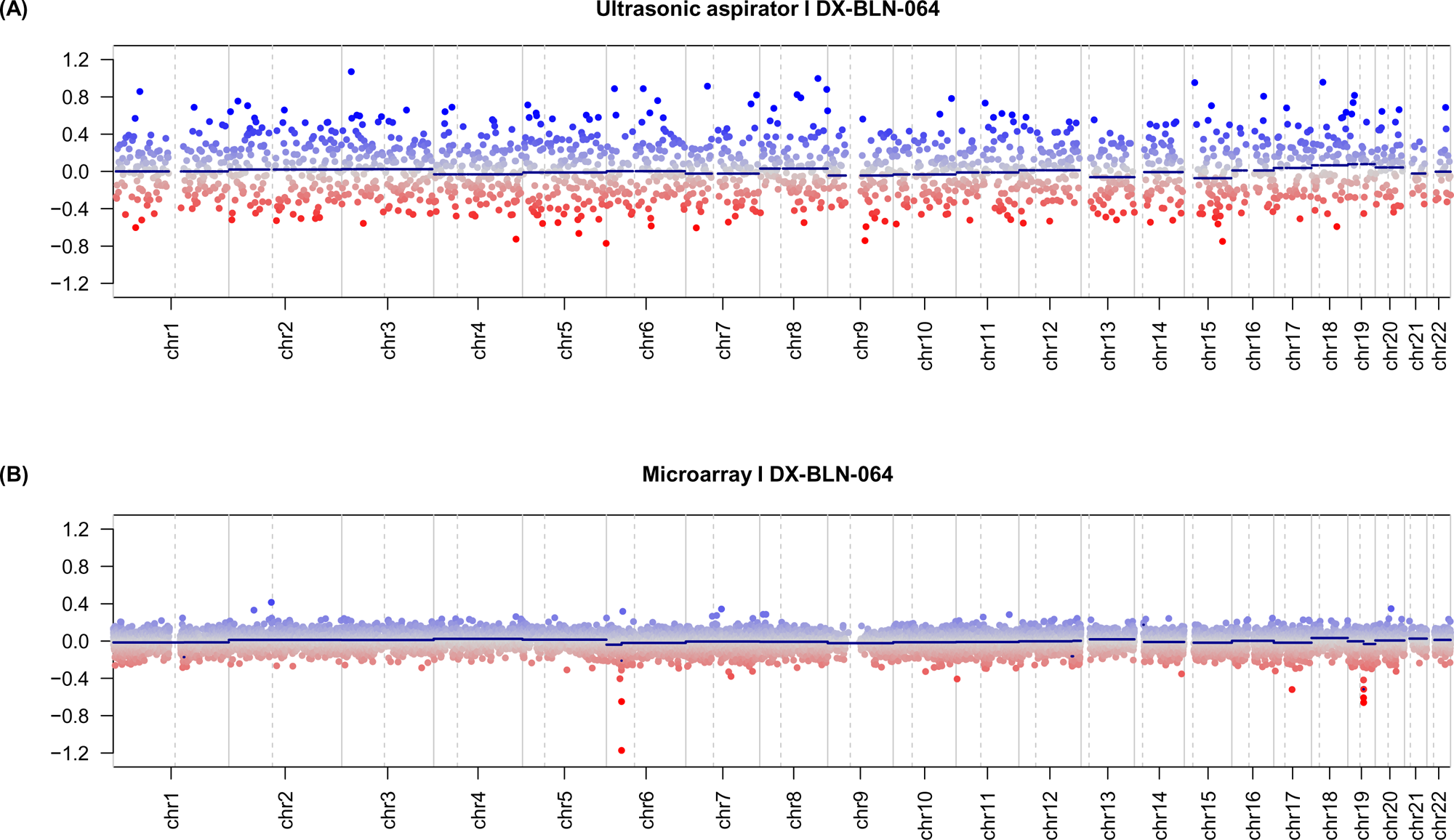
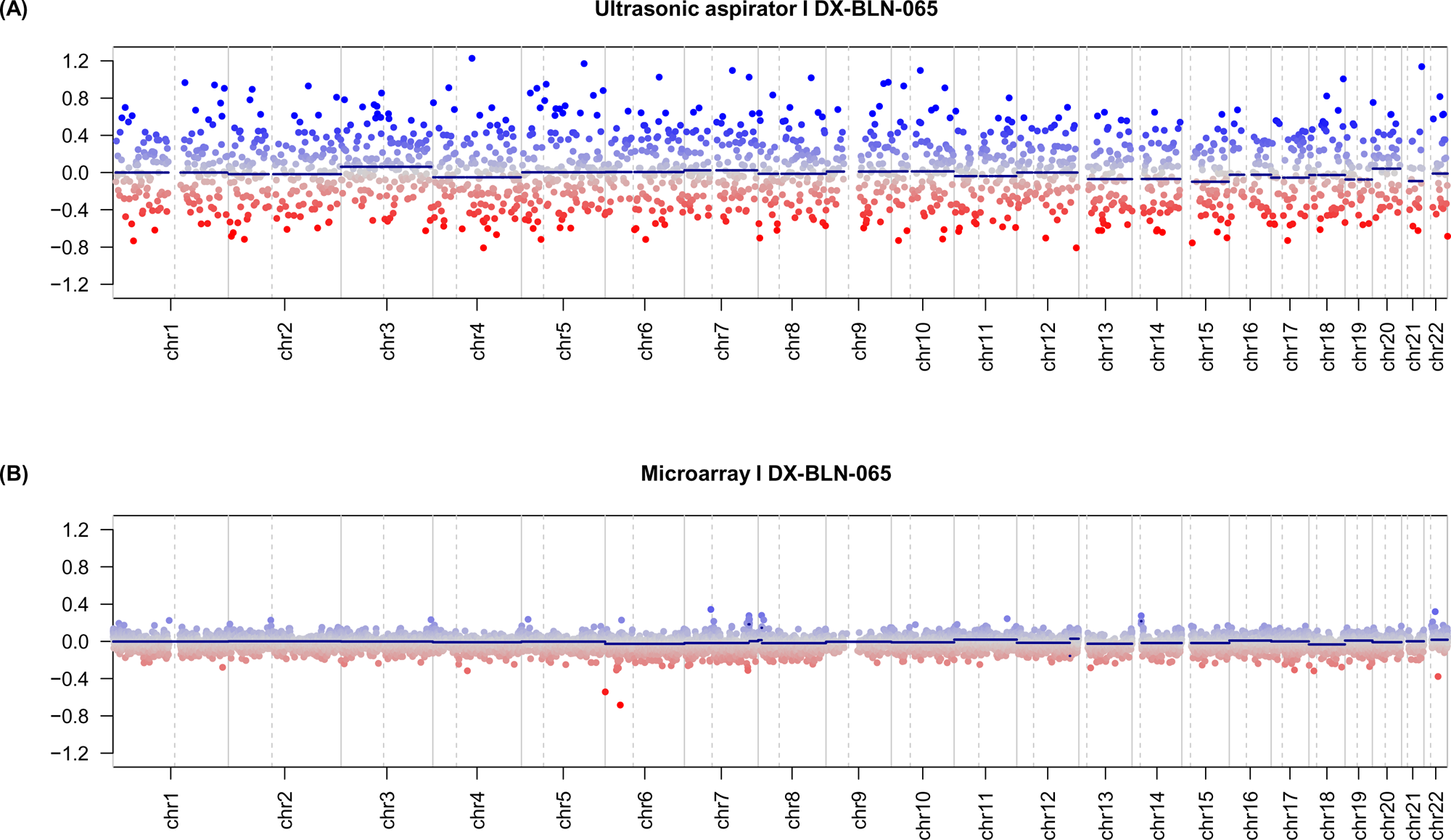
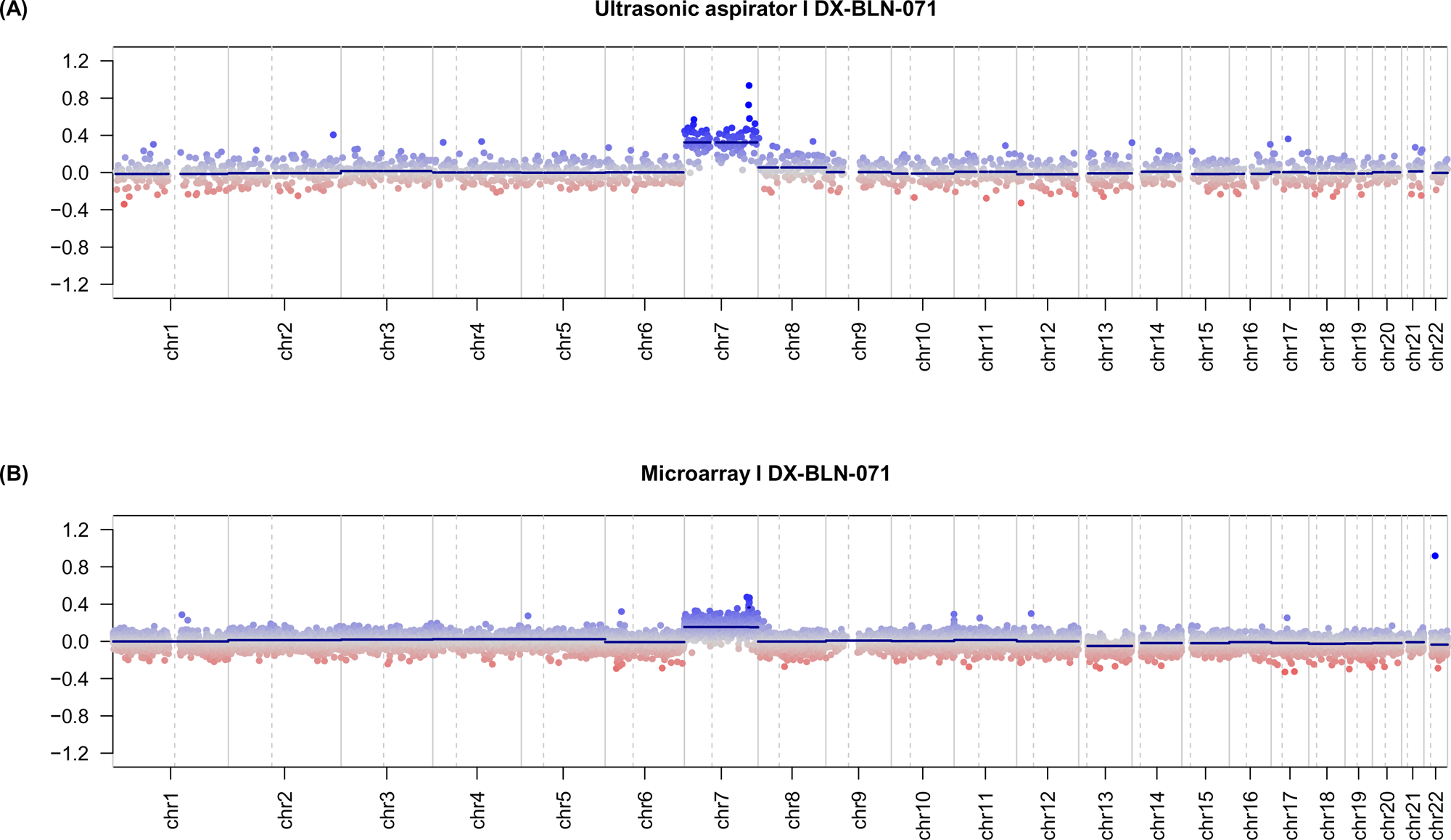
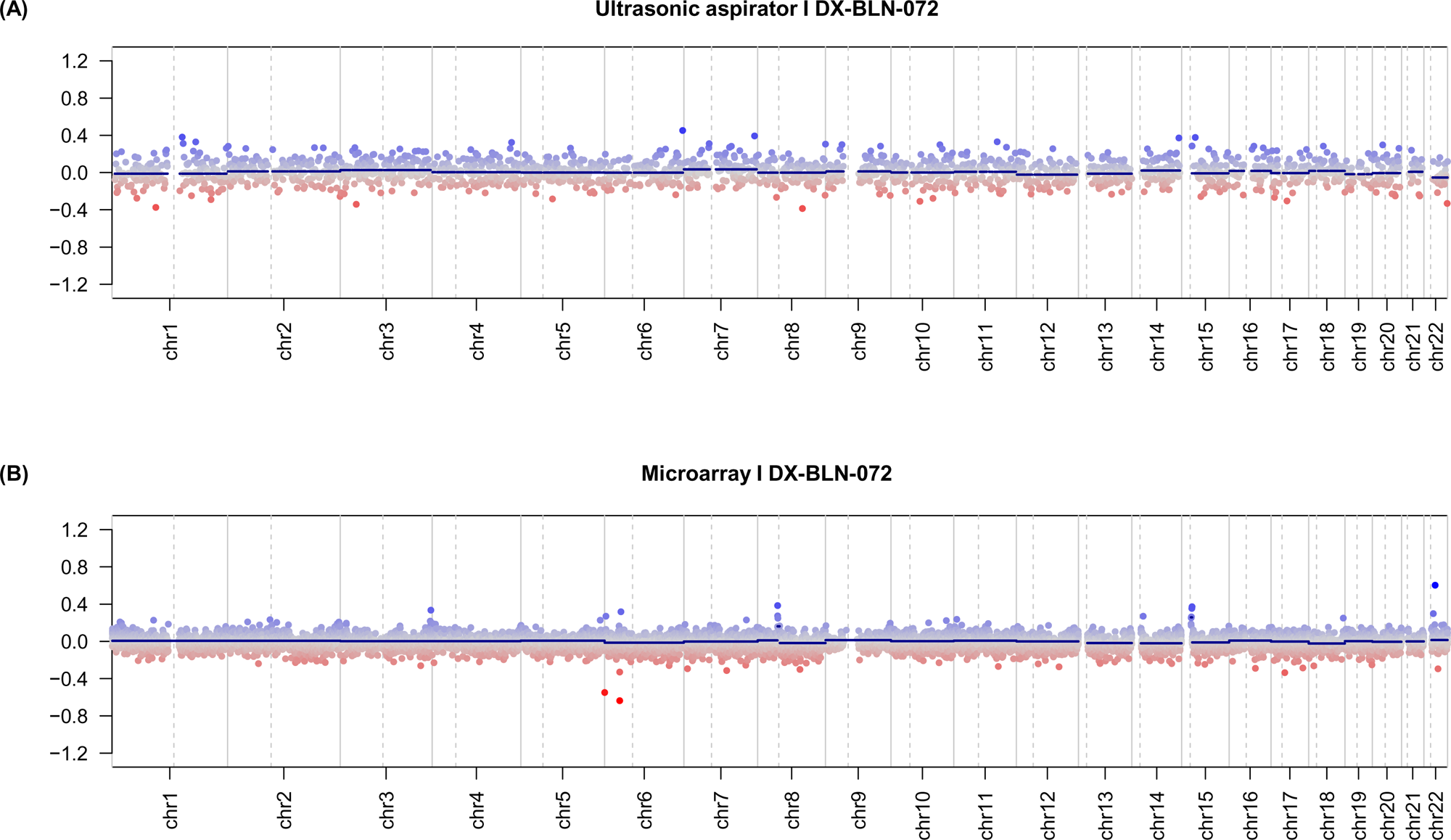
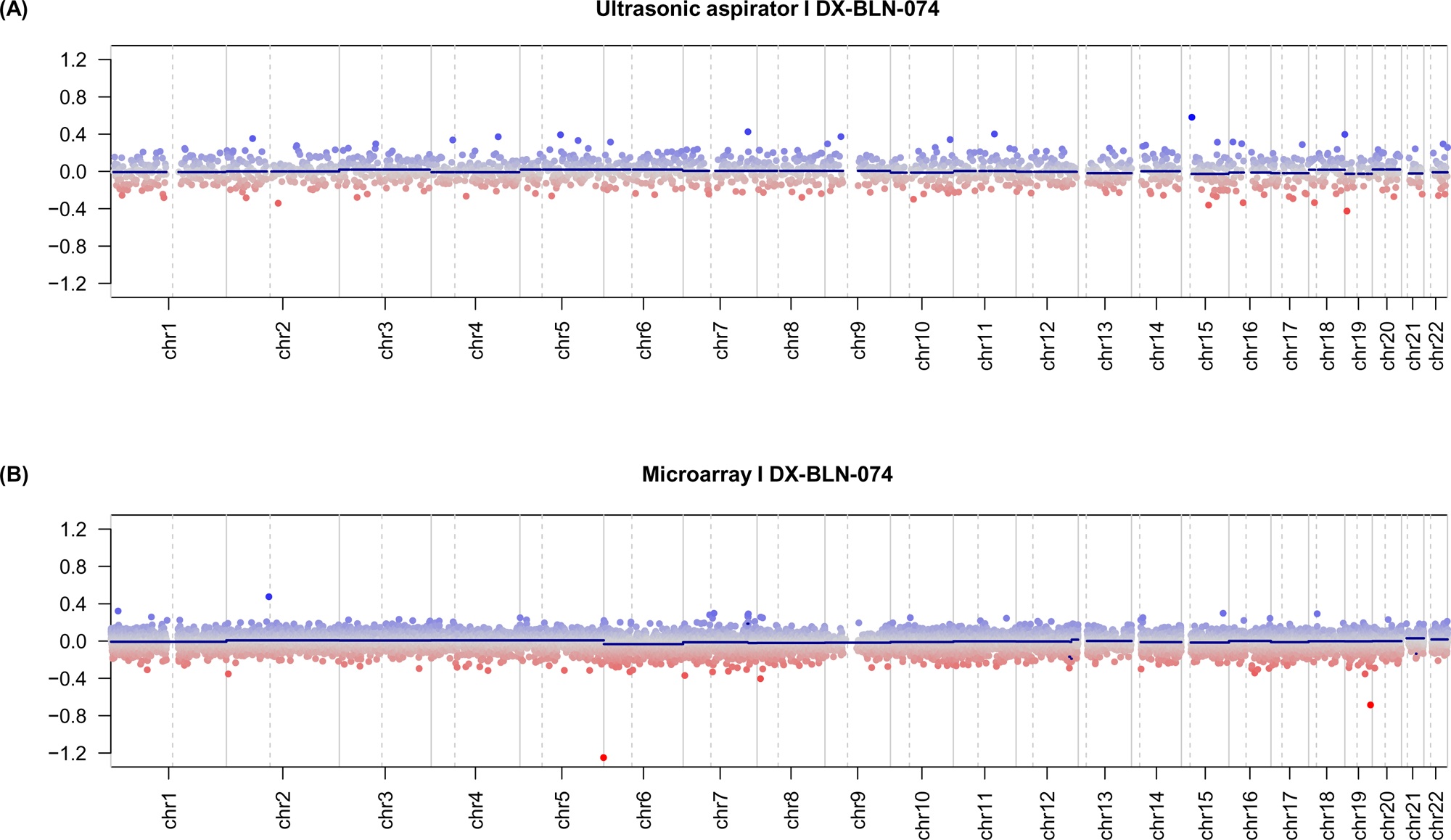
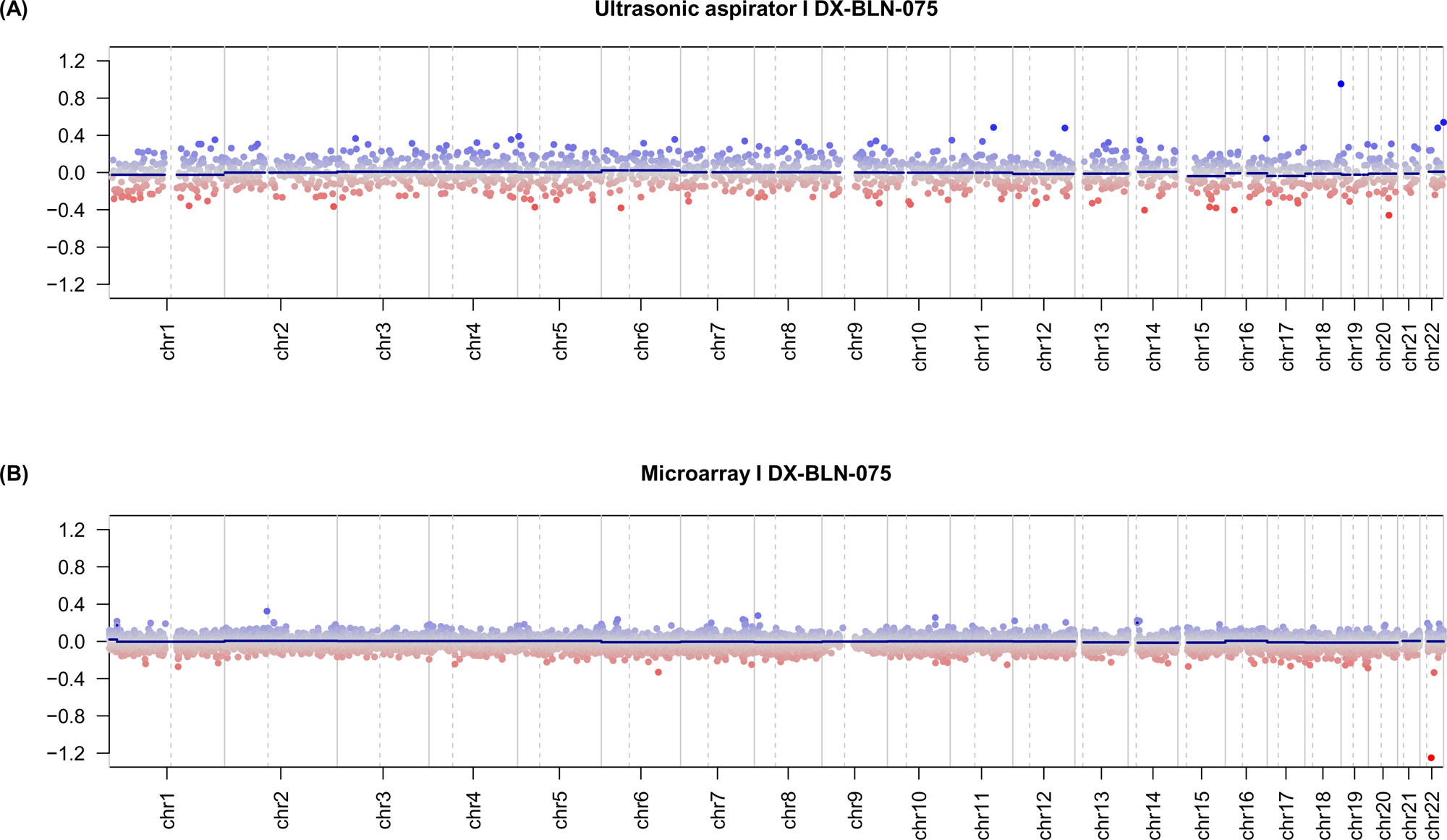
Comparison of copy number variation profiles obtained from **(A)** ultrasonic aspirator tissue samples and nanopore sequencing and **(B)** FFPE tumor tissue subjected to EPIC microarray (850K).

